# Improved performance of nucleic acid-based assays for genetically diverse norovirus surveillance

**DOI:** 10.1101/2023.03.13.23286721

**Authors:** Chamteut Oh, Aijia Zhou, Kate O’Brien, Arthur R Schmidt, Joanna L. Shisler, Arthur R Schmidt, Laura Keefer, William M. Brown, Thanh H. Nguyen

## Abstract

Nucleic acid-based assays, such as polymerase chain reaction (PCR), that amplify and detect organism-specific genome sequences are a standard method for infectious disease surveillance. However, challenges arise for virus surveillance because of their genetic diversity. Here, we calculated the variability of nucleotides within the genomes of ten human viral species *in silico* and found that endemic viruses exhibit a high percentage of variable nucleotides (e.g., 51.4% for norovirus GII). This genetic diversity led to variable probability of detection of PCR assays (the proportion of viral sequences that contain the assay’s target sequences divided by the total number of viral sequences). We then experimentally confirmed that the probability of the target sequence detection is indicative of the number of mismatches between PCR assays and norovirus genomes. Next, we developed a degenerate PCR assay that detects 97% of known norovirus GII genome sequences and recognized norovirus in eight clinical samples. In contrast, previously developed assays with 31% and 16% probability of detection had 1.1 and 2.5 mismatches on average, respectively, which negatively impacted RNA quantification. Additionally, the two PCR assays with lower probability of detection also resulted in false negatives for wastewater-based epidemiology. Our findings suggest that the probability of detection serves as a simple metric for evaluating nucleic acid-based assays for genetically diverse virus surveillance.

**Importance:** Nucleic acid-based assays, such as polymerase chain reaction (PCR), that amplify and detect organism-specific genome sequences are a standard method for infectious disease surveillance. However, challenges arise for virus surveillance because of the rapid evolution and genetic variation of viruses. The study analyzed clinical and wastewater samples using multiple PCR assays and found significant performance variation among the PCR assays for genetically diverse norovirus surveillance. This finding suggests that some PCR assays may miss detecting certain virus strains, leading to a compromise in detection sensitivity. To address this issue, we propose a metric called the probability of detection, which can be simply calculated *in silico* using a code developed in this study, to evaluate nucleic acid-based assays for genetically diverse virus surveillance. This new approach can help improve the sensitivity and accuracy of virus detection, which is crucial for effective infectious disease surveillance and control.

## Introduction

Tracking pathogens is a fundamental public health intervention to control communicable diseases (1). Nucleic acid-based assays that analyze organism-specific genome sequences, such as polymerase chain reaction (PCR), loop-mediated isothermal amplification (LAMP), recombinase polymerase amplification (RPA), and clustered regularly interspaced short palindromic repeats (CRISPR)-based assays (2), are employed widely as a standard method for virus surveillance. However, challenges arise in virus surveillance utilizing these nucleic acid-based assays because virus genomes constantly mutate (3). The viral genome mutation rate is typically orders of magnitude faster than that of the other types of microorganisms, such as bacteria, because viral RNA polymerases normally lack genome proofreading and repair activities (4, 5). This high mutation rate enables viruses to evolve quickly, resulting in genetic variation within the virus population (6). For instance, norovirus is composed of ten genetically variable genogroups (from GI to GX) (7), rotavirus has been reported to have eight distinct strains (from A to H) (8), and human adenovirus is subdivided across seven separate species (from A to G) (9). As the viral sequence is a crucial determinant for the sensitivity and specificity of nucleic acid-based assays (10), genome diversity and stability must be considered in the design of nucleic acid-based assays for viruses.

The design of nucleic acid-based assay reagents, such as primers and probes for PCR, typically involves two stages: *in silico* sequence analysis and *in vitro* verification. For example, six human rotavirus sequences (G2P4, G3P14, G8P6, G12P6, and G1P8) were used to design primer and probe sequences (11). The authors then used 121 human fecal samples collected from 2004 to 2006 in Slovenia to verify their PCR assays. Another study used 178 human rotavirus A VP6 gene sequences to design primers and a probe (12). The authors then used 266 human samples collected between 2009 and 2012 from Western India for assay verification. Likewise, 18 human rotavirus sequences and 775 clinical samples collected between 2010 and 2014 in Sweden (13). Because these reverse transcriptase qPCR (RT-qPCR) assays were designed with different rotavirus sequences and verified with different clinical samples, they may not show the same performance in a certain scenario, for example, where rotavirus surveillance is conducted in the USA in 2023. Indeed, there is a lack of research evaluating the impacts of different nucleic acid-based assays on virus surveillance.

We hypothesize that a nucleic acid-based assay targeting a specific genome sequence would show different genome amplification efficiencies based on the genome diversity of a virus population, and this would ultimately affect virus surveillance. To test this hypothesis, we first carried out *in silico* analysis to determine the level of nucleotide diversity across the genomes of ten viral species. Our results showed that endemic virus surveillance could be significantly impacted by the target sequences of nucleic acid-based assays. We then conducted *in vitro* experiments using multiple RT-qPCR assays for norovirus on clinical and environmental samples. These experiments revealed that an RT-qPCR assay could exhibit a substantially reduced amplification efficiency when compared to other assays, depending on the norovirus genotypes. Based on these findings, we propose a straightforward approach for evaluating the performance of nucleic acid-based assays for virus surveillance in advance.

## Materials and Methods

### Viral genetic diversity analysis

We obtained complete viral sequences from open-source databases, such as Genbank (ncbi.nlm.nih.gov/genbank) or Global Initiative on Sharing All Influenza Data (GISAID; gisaid.org) to investigate the genetic diversity of viruses. We downloaded all available viral sequences from the databases and, in cases where sufficient sequences were available, we applied an additional filter of collection location and year to complete *in silico* analysis using an ordinary laptop. The viral sequences were aligned using MUSCLE (v3.8.1551) (14). To expedite the computation, we sliced the complete genome into smaller pieces for viruses with a long sequence length and/or numerous sequences. We compared sequences between the complete alignment and individual viral sequences using Jalview (v.2.11.2.5) (15).

We defined nucleotide identity as the percentage of the consensus nucleotide of the alignment. When we calculated nucleotide identity using Jalview (16), gaps in each viral sequence that occurred during the genome sequencing process (i.e., undetermined sequence) or were due to different sequence lengths, were ignored. Additionally, if a consensus nucleotide was determined with less than ten viral sequences, those nucleotides were excluded from the alignment. We defined nucleotide identity lower than 90% as a variable nucleotide, and used it to evaluate the level of genome diversity. Further details about sequence download and alignment determination can be found in **Supporting Information**.

### *In silico* probability of target sequence detection of (RT-)qPCR assays

We determined the impact of viral genome diversity on the performance of (RT-)qPCR by calculating the probability of detection. We used an algorithm (17) to calculate the probability of detection, which is the ratio of the number of viral genomes that have identical sequences to those of the (RT-)qPCR assay (e.g., sequences of two primers and one probe) to the total number of viral genomes found database, such as Genbank or GISAID. Then, we used the probability of detection to evaluate the sensitivity and specificity of the SASR-CoV-2 variant-specific PCR assays. For this study, we improved the algorithm, which has been published previously, to calculate the probability of detection of (RT-)qPCR assays with degenerate sequences (https://github.com/Nguyen205/In-silico-analysis-for-degenerate-qPCR-assay). The input data for calculating probability of detection of the (RT-)qPCR assays are viral sequences in a fasta format obtained from the database and sequences of (RT-)qPCR assay.

### Degenerate RT-qPCR assay design to detect genetically variable norovirus populations

We collected 637 complete norovirus sequences from Genbank and determined their genogroups and genotypes using the Norovirus Genotyping Tool Version 2.0 (www.rivm.nl/mpf/typingtool/norovirus). The sequences whose genotypes were not determined with this tool were excluded from further analysis. Although noroviruses are genetically diverse, including ten genogroups (7), the majority of norovirus infections have been caused by the GI and GII genogroups (18, 19). We also found that most of our sequences were either GI (n=48) or GII (n=561), while the other genogroups were negligible, showing only three sequences of GIII and eight sequences of GIV. For this reason, we designed two RT-qPCR assays, each targeting GI and GII genogroups, respectively.

Next, we aligned the sequences of each genogroup with MUSCLE (v3.8.1551) (14). The aligned sequences were used to generate degenerate sequences, which contains multiple possible nucleotides at one position, using the DegePrime (20). We selected degenerate sequences that were longer than 18 bases for a primer and 20 bases for a probe, had a degeneracy of less than 24, and an amplicon size between 80 and 200 bases. We further analyzed these degenerate sequences using Oligoanalyzer (Integrated DNA Technologies, USA) to ensure that they met the following requirements. First, the degenerate primers and probes must not have degenerate nucleotides at the last three bases from 3’ end. Second, the average probes’ melting temperature (T_m_) must be 7°C higher than those of primers. Finally, the probe sequence must not start with a guanine from the 5’ end.

Once the candidate primer and probe sequences that satisfied all of the requirements outlined above were determined through *in silico* analysis, we conducted an *in vitro* experiment to evaluate the performance of these primers and probes in terms of amplification of the target sequences. These oligonucleotides were synthesized by Integrated DNA Technologies (USA). First, we analyzed melting curves from SYBR-based one-step RT-qPCR analysis and did not detect any obvious evidence for the formation of primer-dimers (**Fig. S1**). Second, we generated calibration curves with synthetic DNA controls (Integrated DNA Technologies, USA) (**Table S1**) to confirm that the PCR efficiency fell within the range from 85% to 110% and that the R^2^ value was greater than 0.99. Third, we determined the limit of detection (LOD) for the RT-qPCR assay developed in this study, using 20 replicates of serial dilutions of synthetic controls, following a previous study (17). We found that the LODs for GI and GII were 1.1 and 5.7 gc/μL, respectively (**Fig. S2**). Fourth, we evaluated the specificity of the RT-qPCR to our target sequences. This is an important step, as we designed degenerate PCR assays to cover a wide range of norovirus genotypes. We used clinical samples of GI to test the specificity of the GII assays and vice versa, as these samples contain a high concentration of microbes from humans, including norovirus GI (which is expected to have higher sequence similarity to GII samples). We did not detect fluorescence signals from GI clinical samples when we used GII assays, or vice versa (**Fig. S3**). Furthermore, *in silico* analysis showed that the GI and GII assays had 0% of probability of detection for GII (n=561) and GI viral sequences (n=48), respectively. These results support that our GI and GII assays specifically amplify target viral sequences. We chose the final primers and probes that showed the highest probability of detection among the candidates, satisfying all *in silico* and *in vitro* verification steps (**Table S1**).

### RT-qPCR protocol for norovirus quantification

The SYBR-based RT-qPCR analysis was conducted to detect the formation of primer-dimers. The RT-qPCR mixture for SYBR-based RT-qPCR assay included 3 μL of RNA sample, 0.3 μL of 50 μM forward and reverse primer, 1.275 μL of molecular biology grade water (Corning, NY, USA), 5 μL of 2×iTaq universal SYBR green reaction mix, and 0.125 μL of iScript reverse transcriptase from the iTaq™ Universal SYBR® Green One-Step Kit (1725151, Bio-Rad Laboratories, USA). The PCR cocktail was placed in 96-well plates (4306737, Applied Biosystems, USA) and analyzed by an RT-qPCR system (QuantStudio 3, Thermo Fisher Scientific, USA). The RT-qPCR reaction was performed with a thermocycle of 50°C for 10 minutes and 95°C for 1 minute, and then 40 cycles of denaturation at 95°C for 10 seconds, annealing at 53°C for 30 seconds, and extension at 60°C for 30 seconds. Melting curves were analyzed while the temperature increased from 60°C to 95°C. The SYBR signal was normalized to the ROX reference dye. The cycles of quantification (Cq) were determined by QuantStudio Design & Analysis Software (v1.5.1).

The Taqman-based RT-qPCR assays were conducted for genome quantification. The Taqman-based RT-qPCR started by mixing 2.5 μL of RNA sample, 2.5 μL of Taqman Fast Virus 1-step Master Mix (4444432, Applied Biosystems, USA), and 5 μL of primers/probe mixture to achieve final concentrations of 2000 nM for primers and 1000 nM for probes. The PCR cocktail was placed in 96-well plates (4306737, Applied Biosystems, USA) and analyzed by an RT-qPCR system (QuantStudio 3, Thermo Fisher Scientific, USA) with a thermal cycle of 5 minutes at 50°C, 20 seconds at 95°C followed by 40 cycles of denaturation at 95°C for 15 seconds, annealing at 53°C for 30 seconds, and extension at 60°C for 30 seconds. When previously designed RT-qPCR assays were used, the annealing temperature was adjusted for each RT-qPCR assay as reported in references (**Table S1**).

For both SYBR- and Taqman-based assays, at least three replicates were analyzed for serial dilution of synthetic DNA (for a standard curve), nuclease-free water (as a negative control), and samples. All positive samples were positive and negative samples were negative in all RT-qPCR analyses. The linear dynamic range for the serial dilutions of synthetic DNA was between 10^0^ to 10^5^ gc/uL. The PCR efficiencies for RT-qPCR were higher than 85% (R^2^>0.99). The details for RT-qPCR assays are summarized in **Table S2**, in accordance with MIQE guidelines (10).

### Clinical samples collection and processing

Twenty unidentified stool samples collected from norovirus-infected patients were provided by the Illinois Department of Public Health. Sample collection dates and locations are summarized in **Table S3**. Ten samples were positive for GI genogroup, and the other ten samples were for GII genogroup. Norovirus RNA was extracted using the following procedure. One hundred milligrams of stool sample were mixed with 900 μL of DI water. The mixtures were vortexed for 30 seconds and centrifuged at 17,000g for 10 minutes. Viral RNA was extracted from 140 μL of the supernatant using QIAamp Viral RNA mini kit (Qiagen, Germany) following the manufacturer’s protocol (21–23). An inhibition test was conducted by adding Tulane virus (whose host is a rhesus monkey) to each extract, following our previous protocol (24). We found the impact of any possible inhibitors was negligible (**Fig. S4**). The RNA extracts were kept at -80°C until downstream analysis. Clinical samples were analyzed by Sanger sequencing to obtain viral sequences (**Supporting Information, Table S4** and **S5**).

### Sewage sample collection and processing

We collected wastewater samples from a city-scale and a neighborhood-scale sewershed. Specifically, we obtained influent wastewater from the Urbana-Champaign Sanitary District (IL, USA), which serves 144,097 people living in Champaign city, Urbana city, and adjacent areas from January 2022 to May 2022. In addition, we collected samples from a manhole receiving wastewater discharged by 1675 residents from January 2021 to May 2022. All wastewater samples were obtained using an autosampler (Teledyne ISCO, USA), programmed to collect a 1 to 2 L composite sample comprised of samples pumped for 24 hours. The composite samples were transferred to sterile sampling bags (14-955-001, Fisher Scientific, USA), and 20 mL of 2.5 M MgCl_2_ was added to the samples (i.e., final MgCl_2_ concentrations were from 25 to 50 mM) to coagulate solids including virus particles (24, 25). The samples were transported on ice to a laboratory at the University of Illinois Urbana-Champaign within three hours. Upon arrival at the laboratory, supernatants from each composite sample were discarded. The remaining 35 mL of sewage, in which solid particles were concentrated, were transferred to a 50 mL tube (12-565-271, Fisher Scientific, USA). The sewage samples were centrifuged at 10,000 g for 30 minutes (Sorvall™ RC 6 Plus, Thermo Scientific, USA). Supernatants were discarded, and a portion of the concentrated sludge (100 μL) was transferred to a sterile 1.5 mL tube (1415-2600, USA Scientific, USA). Nucleic acids were extracted from the sludge with QIAamp Viral RNA mini kit (Qiagen, Germany) following the manufacturer’s procedure. Sewage collection and processing were conducted on the same day, and the RNA samples were stored at -80°C until RT-qPCR analysis. RNA quantification was conducted with a 10-fold dilution of RNA extracts to lower the impact of the inhibitors to a negligible level (24).

### Statistical analysis

Mann-Whitney U test was conducted to compare the nucleotide identity of two viral species (**Fig. 1**). Linear regression analysis was conducted to compare norovirus RNA concentrations of clinical samples determined by two RT-qPCR assays (**Fig. 2**). The slope of the linear regression curve was compared to 1. Samples with a studentized residual of less than -1.5 were defined as outliers and excluded from the linear regression curve to evaluate the potential impacts of mismatch between RT-qPCR assays and norovirus sequences (**Fig. 2**). Statistical analyses were conducted using OriginPro 2023.

**Fig. 1.**
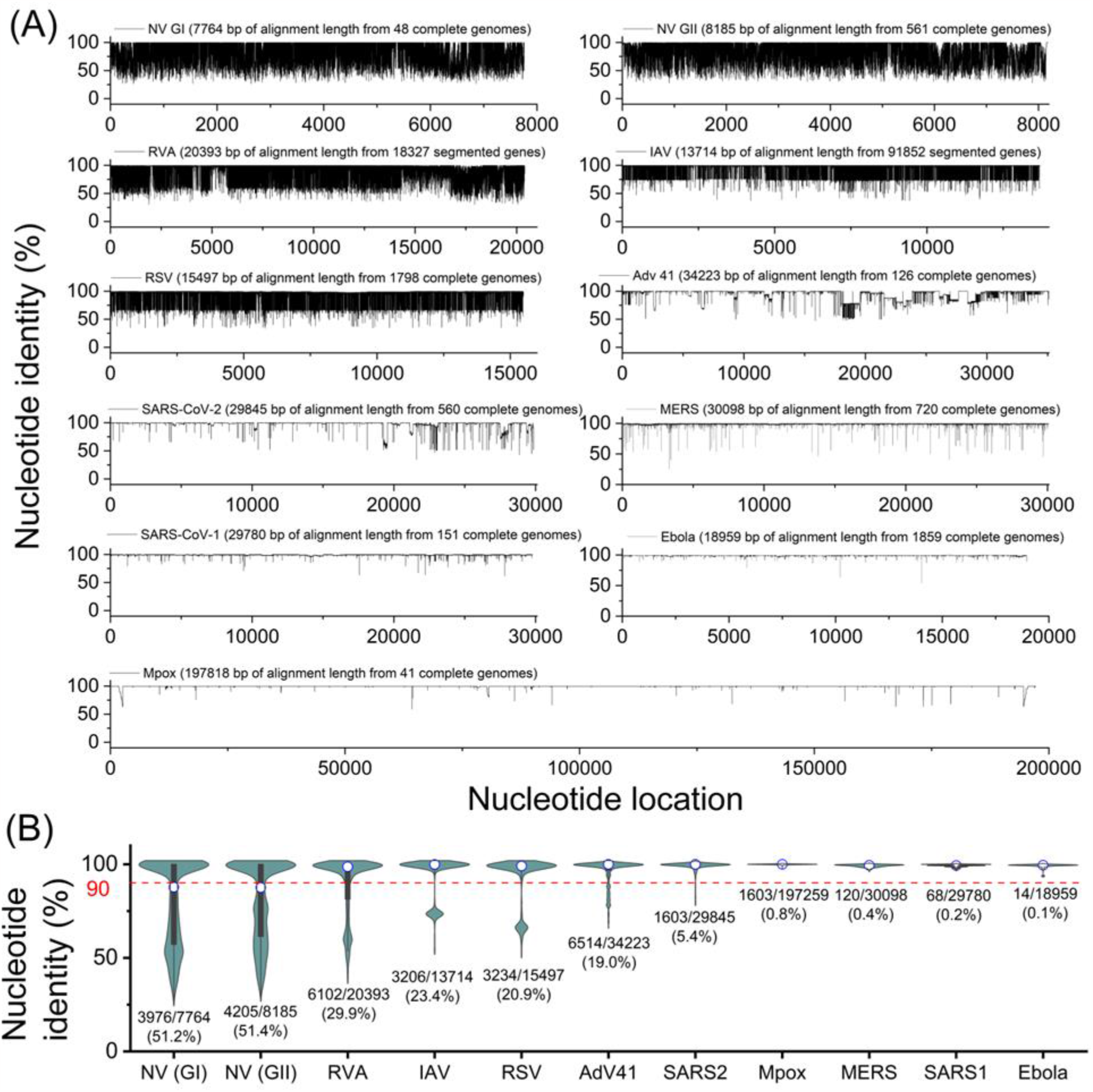
Genetic diversity analysis of ten viral species. **(A)** Percent nucleotide identity of the alignment for each viral species with nucleotide locations. **(B)** Summary of Nucleotide identity in bar charts for each viral species. The figures below each bar chart indicate the number of variable nucleotides/the number of total nucleotides (a percentage of variable nucleotides). Any two viral species showed significantly different nucleotide identities (Mann-Whitney U test, p<0.05).

**Fig. 2.**
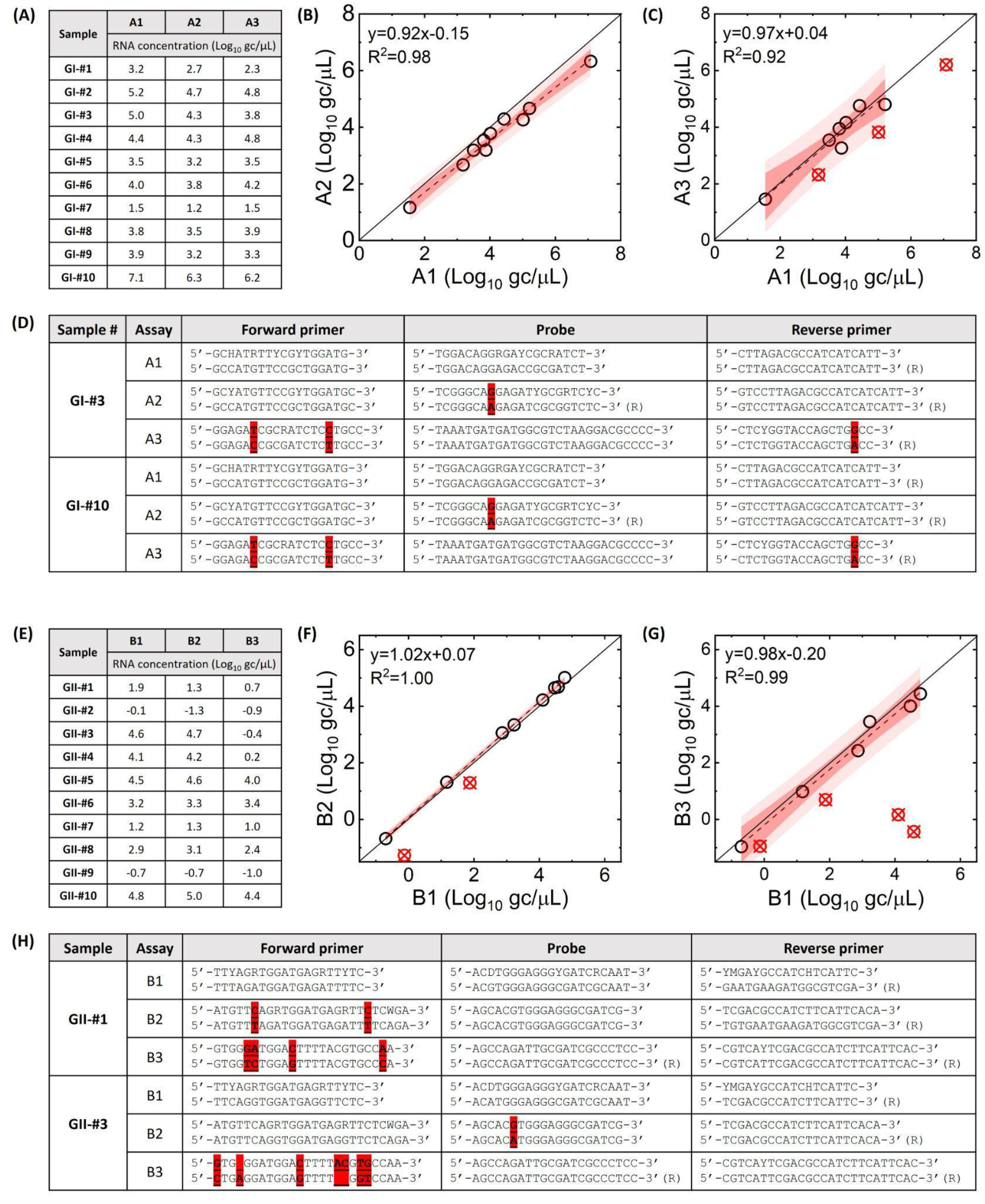
Clinical samples analyzed by RT-qPCR assays. **(A)** and **(E)** show GI and GII RNA concentrations, respectively. GI RNA concentrations by A1 assay are compared to those by A2 **(B)** and A3 assay **(C)**. GII RNA concentrations by B1 assay are compared to those by B2 **(F)** and B3 assay **(G)**. Red circles with a cross indicate outliers that deviated from the regression line (a cut-off studentized residual is -1.5). Dark and pale shades represent 95% confidence and prediction intervals, respectively. Possible annealing sequences for GI-#3 and GI-#10 with A1, A2, and A3 assays are presented in **(D)**. Possible annealing sequences for GI-#1 and GI-#3 with B1, B2, and B3 assays are presented in **(H)**. Sequences of RT-qPCR assay and viral genome are illustrated top and bottom, respectively. Highlighted nucleotides show mismatches.

## Results

### Viral genetic variability leads to a wide range of probability of detection of PCR assays

We analyzed genome sequences of ten viral species, including norovirus (NV), rotavirus (RV), respiratory syncytial virus (RSV), influenza virus (IV), adenovirus (AdV), severe acute respiratory syndrome coronavirus 1 and 2 (SARS-CoV-1 and SARS-CoV-2, respectively), middle east respiratory syndrome (MERS), Ebola (which caused the 2013 outbreak), and monkeypox virus (Mpox; which caused the 2022 outbreak), to cover a wide range of viral genomic characteristics from an evolutionary perspective. For example, NV, RV, RSV, IV, SARS-CoV-1, SARS-CoV-2, MERS, and Ebola are RNA viruses, while AdV and Mpox are DNA viruses. These viruses also caused zoonotic outbreaks in humans at different times. Mpox has been causing outbreaks in humans for decades, most recently in 2022. SARS-CoV-1 caused human disease in 2002, while MERS and Ebola viruses caused human outbreaks in 2012 and 2013, and SARS-CoV-2 started infecting humans in 2019, while the other viruses were reported earlier and have been causing outbreaks.

We compared a group of viral sequences to their alignment sequence and determined nucleotide identity (i.e., the percentage of the consensus nucleotide) for the whole viral genomes in **Fig. 1A**. As viruses mutate and the mutations are passed on to subsequent generations, the viral genomic sequence becomes increasingly diverse, and the nucleotide identity decreases (26, 27). Thus, nucleotide identity has been used to describe genomic diversity (28, 29). In this study, we defined a variable nucleotide as a nucleotide with less than 90% of nucleotide identity to evaluate the level of genome diversity. We found that variable nucleotides, below the red lines in **Fig. 1A**, are distributed throughout the entire viral genomes. Next, we summarized the nucleotide identity for each viral species in a bar chart and determined the percent variable nucleotides that vary substantially depending on viral species (**Fig. 1B**). We found the percentage of variable nucleotides. Specifically, NV (51.2% for GI and 51.4% for GII), RV (29.9%), IV (23.4%), RSV (20.9%), and AdV (19.0%) contain more than 10% variable nucleotides. In contrast, SARS-CoV-2 (5.3%), Mpox (0.8%), MERS (0.4%), SARS-CoV-1 (0.2%), and Ebola (0.1%) represented relatively low values of variable nucleotide. Note that the percentage of variable nucleotides was calculated using only one strain for some viral species, including NV (GI and GII genogroups), RV (rotavirus A strain), IV (influenza A virus), AdV (adenovirus type 41) because the alignment with all strains could not be created due to the genome complexity. Thus, the actual variable nucleotides for these viral species are likely higher than the values in **Fig. 1**. Interestingly, despite the unprecedented COVID-19 pandemic, which has infected more than 673 million cases globally as of February 2023 since the first case was confirmed in 2019, the variable nucleotides of SARS-CoV-2 were fewer than the endemic viruses that were reported earlier and have caused outbreaks.

The high level of genetic diversity for the endemic viruses suggests that it may be challenging to find a conserved region that can be targeted by nucleic acid-based assay, such as (RT-)qPCR.

We conducted *in silico* analysis to calculate the probability of detection that previously published primer sets for (RT-)qPCR assays would detect these genome variants (**Tables S1** and **S6**). We found that the probability of detection significantly varied depending on RT-qPCR assays. For example, RT-qPCR assays for RSV designed by three separate studies only detected 2%, 23%, and 46% of known RSV variants, respectively, when applied to sequences we obtained from Genbank (30–32). Similarly, qPCR assays for AdV type 41 by three groups present probabilities of 43%, 77%, and 100%, respectively, for sequences we obtained from Genbank (33–35). The probability of detection for RV by other three studies were calculated to be 3%, 16%, and 81%, respectively (11, 12, 36). The variations in probability of detection observed in **Tables S1** and **S6** indicate that the (RT-)qPCR assays targeting different viral sequences may present significantly different probability of detection. This finding suggests that different (RT-)qPCR assays could miss the presence of viral variants, which would negatively impact virus surveillance efforts.

### Application of RT-qPCR assays to clinical samples reveals a failure of virus detection by an assay with low probability of detection

We hypothesized that probability of detection for target viral species or strains could serve as an indicator of nucleic acid-based assay performance in virus surveillance. To validate this hypothesis, we first prepared RT-qPCR assays for norovirus with varying levels of probability of detection. We designed RT-qPCR assays with a high probability of detection; the A1 assay with a 100% probability of detection for GI genogroup and the B1 assay with a 97% probability of detection for GII genogroup. Other RT-qPCR assays, such as the A2 (56%) and A3 (8%) for GI and B2 (31%) and B3 (16%) for GII, were adopted from previously published studies (**Table S1**).

Norovirus was selected for the *in vitro* experiment due to its significant genetic variability, as demonstrated in **Fig. 1**, and its importance for public health (37).

The RT-qPCR assays were then applied to clinical samples (**Fig. 2A** and **2E** for GI and GII, respectively). We compared the RNA concentration of the RT-qPCR assays with lower probability of detection levels, such as A2, A3, B2, and B3, to those with high probability, such as A1 and B1 (**Fig. 2B** and **2C** for GI and **Fig. 2F** and **2G** for GII). The comparison results present that some samples (red circle with a cross in **Fig. 2C, F, and G**, respectively) are located significantly below the regression lines (i.e., studentized residual was less than -1.5). These outliers indicate that the RT-qPCR assays with lower detection probability yielded significantly lower RNA concentrations than those with a high detection probability level. For instance, RNA concentrations of GII-#3 and GII-#4 measured by the B3 assay were 10^5.0^- and 10^3.9^-fold lower than those by the B1 assay, respectively (**Fig. 2E**). These results indicate that the B3 assays do not amplify specific norovirus samples as effectively as the B1 assay. Excluding these outliers, slopes of regression analysis among RT-qPCR assays were not significantly different from 1 (p>0.05), meaning that all assays yielded similar RNA concentrations for the rest of the samples.

The reduced amplification efficiencies can be explained by the mismatches between RT-qPCR assay sequences and viral sequences. Potential bindings between primer/probe of RT-qPCR assays and the annealing sites of virus genome are illustrated in **Tables S7-S12, Fig. 2D** and **2H**. We found that GII-#3 and GII-#4, which showed significant reduction in RNA concentration by the B3 assay, had 7 mismatches with the B3 assay while the B1 assay, which had no mismatches with these two samples (**Fig. 2H**). Furthermore, we discovered that the decreased probability of detection by RT-qPCR led to increased number of mismatches between RT-qPCR assays and viral genomes. For example, the A1 assay (100% probability of detection for GI) had no mismatches with the ten GI samples, while the A2 (56% probability of detection) and A3 (13% probability of detection) assays had 0.75 and 2 average mismatches, respectively. Similarly, the B1 assay (97% probability of detection for GII) had no mismatches, whereas the B2 (31% probability of detection) and B3 (16% probability of detection) assays had 1.12 and 2.5 average mismatches, respectively, with the GII samples.

### Mismatches between RT-qPCR assays and norovirus genomes explain RNA quantification of a mixture of norovirus sequences in wastewater

As an important epidemiological tool for disease surveillance, we also evaluated the relationship between detection probability of RT-qPCR assay and results from wastewater-based epidemiology (WBE). We first conducted an experiment in which we spiked local wastewater with known quantities and genotypes of norovirus. In this experiment, we added 2 mg of each clinical sample (i.e., GII-#1, GII-#3, and GII-#10) or mixtures of those samples to 500 mL of local wastewater, from which endogenous norovirus was not detected by the RT-qPCR assays. We then processed the wastewater to obtain the concentrated sludge and quantified norovirus RNA concentrations. As a result, we detected norovirus RNA using the B1 assay, with an average norovirus recovery efficiency of 16.8% (n=7), which is comparable to those by wastewater surveillance procedures for SARS-CoV-2 (38). This finding demonstrates that our sewage processing method can effectively concentrate norovirus RNA from wastewater. We also found that the B2 assay failed to detect GII-#1 and the B3 assay could not amplify GII-#1, GII-#3, and the mixture of these two samples (**Table 1**), which agrees with the results from clinical sample analyses. This finding suggests that variation in RNA concentrations of wastewater among RT-qPCR assays can also be explained by the mismatches between RT-qPCR assays and viral genomes.

**Table 1.**
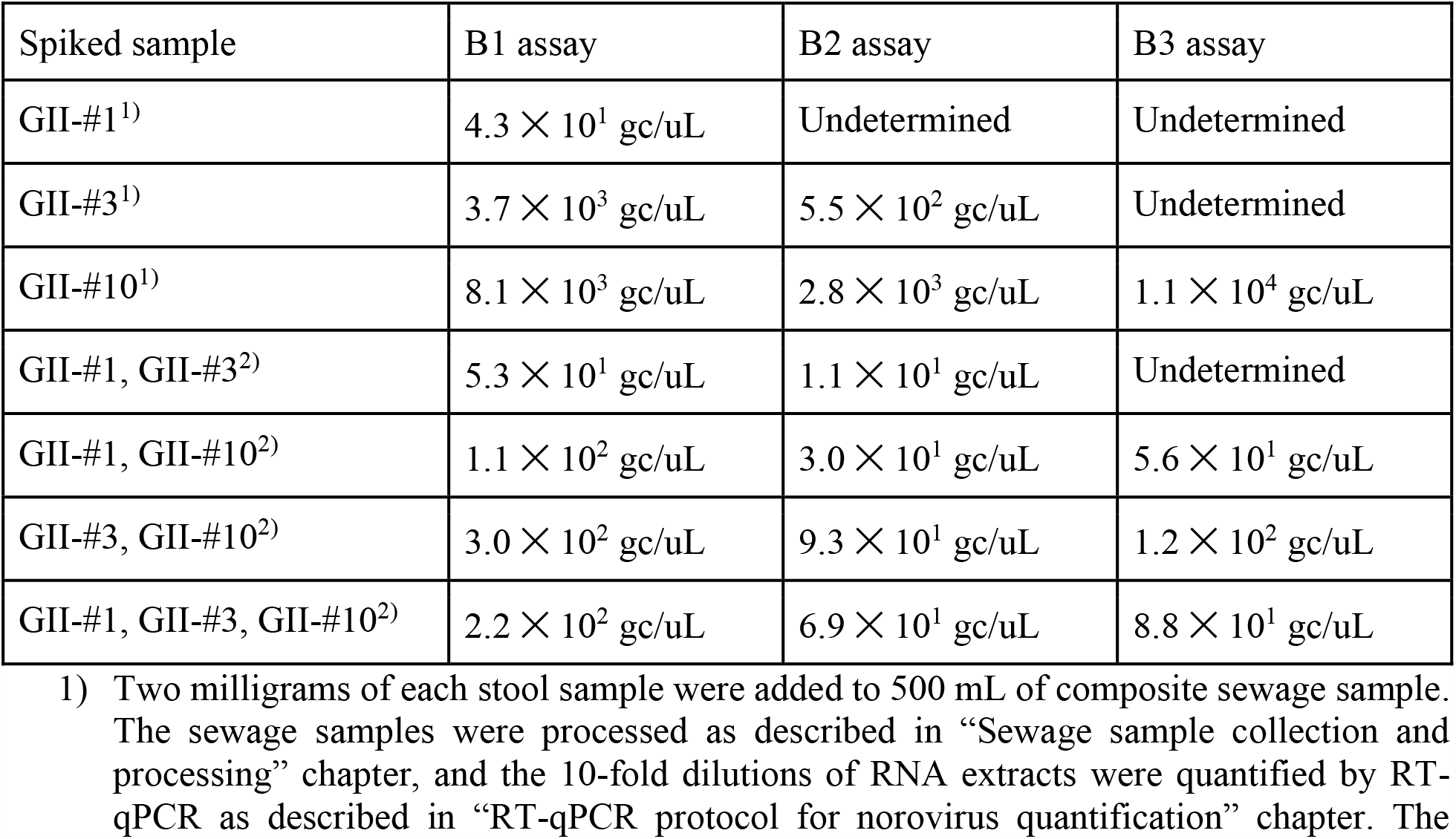
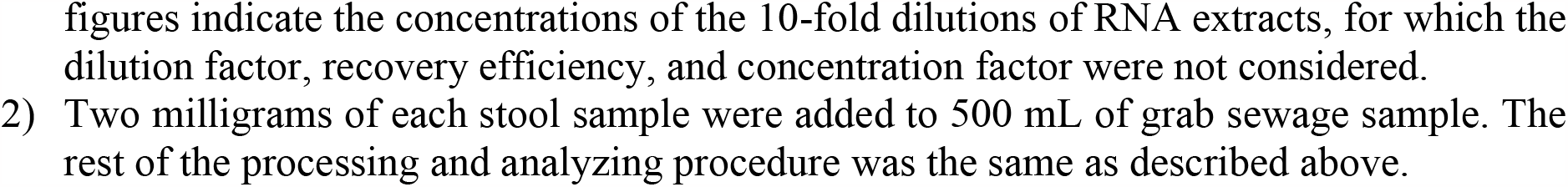
Norovirus RNA concentrations of the norovirus-spiked wastewater quantified by three RT-qPCR assays

### Application of RT-qPCR assays to wastewater samples corroborates the importance of *in silico* analysis for virus surveillance

We utilized RT-qPCR assays to detect norovirus RNA in two sets of wastewater samples collected from city-scale and neighborhood-scale sewersheds (**Fig. 3**). We found that norovirus RNA concentrations of wastewater collected from the city-scale sewershed in 2022 aligned with the percent positive rate of patients by PCR test in Midwestern States (**Fig. S5**), suggesting the norovirus surveillance results were reliable. Interestingly, the RT-qPCR assays showed varying surveillance results depending on the assay probability of detection at a particular monitoring period. For example, from March 2^nd^ to April 21^st^ 2022, the B1 and B2 assays presented a decreasing tendency in GII RNA concentrations, while the B3 assay, with the lowest probability of detection for GII, demonstrated an increasing tendency. Additionally, an RT-qPCR assay with low probability of detection is more susceptible to false negatives. We found that the number of positive samples by A3 and B3 (the lowest probability of detection for GI and GII, respectively) was lower than those by A1 and B1 (the highest probability of detection for GI and GII, respectively) (**Fig. 3E-H**). Thus, we concluded that an RT-qPCR assay with a low probability of detection may result in false negatives in wastewater surveillance.

**Fig. 3.**
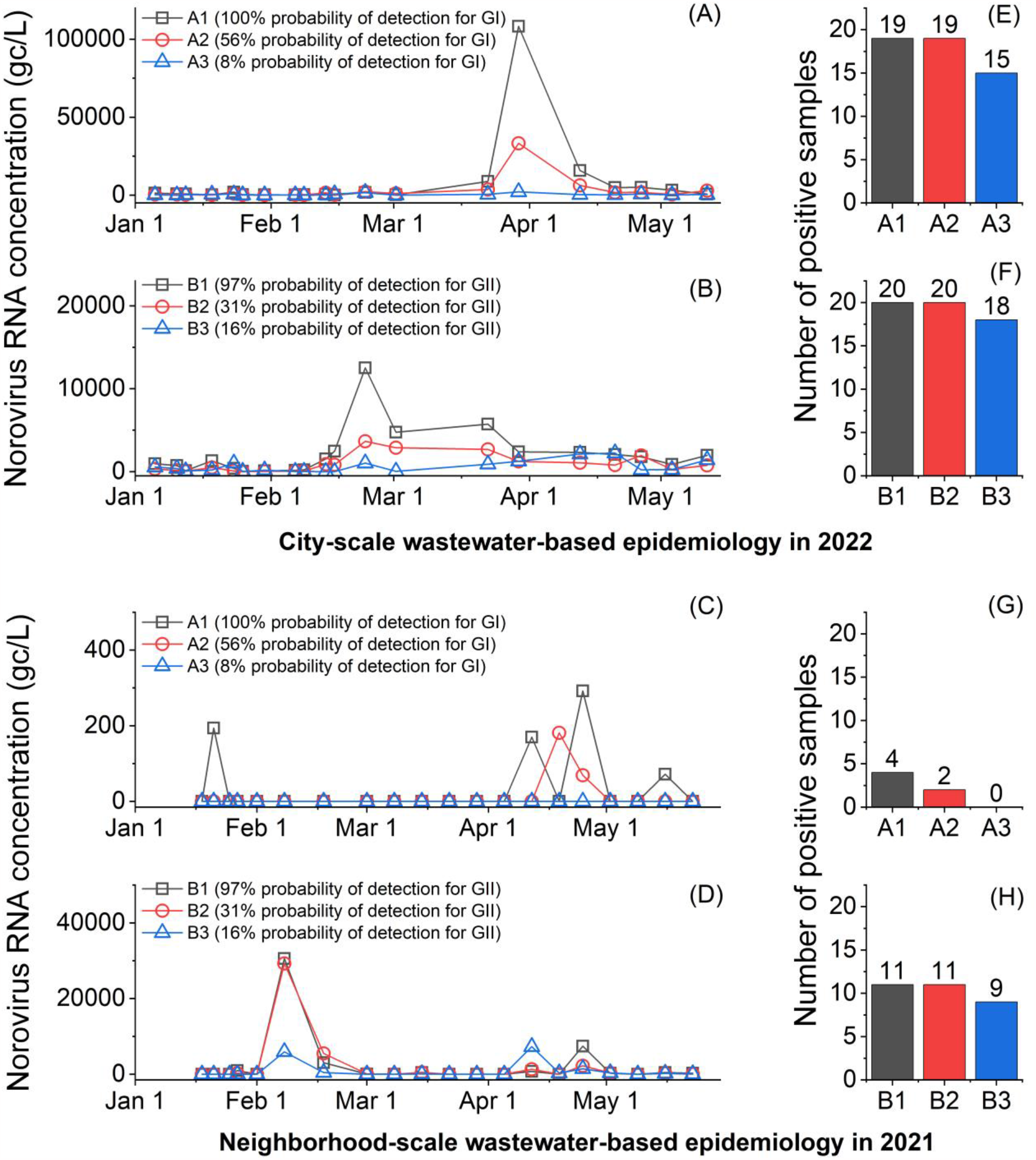
Wastewater samples analyzed by RT-qPCR assays. City-scale and neighborhood-scale wastewater-based epidemiology data are summarized at the top and the bottom, respectively. (A) and (C) present temporal GI concentrations in wastewater, and (B) and (D) show temporal GII concentrations. The numbers of norovirus-positive of wastewater samples are summarized in (E), (F), (G), and (H).

## Discussion

At the emergence of novel viruses in the human population, primarily from zoonotic spillovers from animal reservoirs (e.g., SARS, Ebola, HIV, MERS, Nipah, and Canine parvovirus), their genome sequences are distinct from closely related viruses (39). For instance, bats were identified as natural hosts of coronaviruses closely related to SARS-CoV-1, which caused the 2002–2004 SARS outbreak (40). This bat virus evolved rapidly in at least two intermediate hosts, such as civets, before being transmitted to humans (41), resulting in a SARS-CoV-1 genome that was distinct from previously known groups of coronaviruses (42). If emerging viruses acquire efficient human-to-human transmission, genetic mutations in the viral genomes can be introduced, causing viruses to diverge from their ancestors, as demonstrated in the influenza virus or SARS-CoV-2 variants (43, 44). Depending on disease pathology (mortality, incubation time, transmissibility), and public health interventions (contact tracing, quarantine, and vaccination), some virus strains may fade out and be contained to a limited number of people or even become extinct. On the other hand, other viruses may establish a stable relationship with humans and become endemic viruses, circulating in the human population and intermittently causing outbreaks. Among the viral species in this study, SARS-CoV-2 (45) and Mpox (46) can be considered as newly emerging viruses, while SARS-CoV-1 (47), MERS (48), and Ebola (49) are considered contained viruses, and NV, RV, RSV, IAV, and AdV are classified as the endemic viruses (50).

In this study, we discovered that the endemic viruses exhibited a significantly higher level of genome diversity than the emerging viruses and the contained viruses (Mann-Whitney U test, p<0.05 in **Fig. S5**). This finding can be attributed to the evolutionary rate (i.e., the speed of genetic change in a lineage over a specific period) and the time for which the genetic changes are accumulated (51). **Table S13** summarizes the previously reported evolutionary rates of various viral species. The evolutionary rates ranged from 4 ×10^−4^ to 1.2×10^−2^ nucleotide substitutions/site/year (s/s/y) for the RNA viruses (i.e., TV, RVA, IAV, RSV, SARS, MERS, and SARS-CoV-2) and from 5×10^−6^ to 4.1×10^−5^ s/s/y for the DNA viruses (i.e., AdV and Mpox). Interestingly, AdV, a DNA virus with an evolutionary rate of 4.1×10^−5^ s/s/y, presented a higher degree of genome diversity compared to RNA viruses with faster evolutionary rates, such as SARS-CoV-1 (4 ×10^−4^ s/s/y), SARS-CoV-2 (6.7×10^−4^-3.3 ×10^−3^ s/s/y), MERS (1.1 ×10^−3^ s/s/y), and Ebola (1.2 ×10^−3^ s/s/y). Note that AdV type 41 was estimated to have originated in 1720 (52), allowing the virus with a much longer period for the accumulation of mutations. Although the evolutionary rate reflects the rate at which mutations are passed on to descendants, it is not a direct measure of current genome diversity. Instead, the time for which genetic changes accumulate may play a more critical role. This is why endemic viruses are genetically highly variable.

If a viral species has numerous strains with highly variable genome sequences, many of which are currently circulating in the human population, it is challenging to predict which strains would introduce to a specific location and lead to an outbreak. For example, three geographically adjacent Asian countries suffered from different norovirus genotypes in 2018, showing GII.2 in China (53), GII.4 in Japan (54, 55), and GII.17 in South Korea (56). In this study, we corroborated that RT-qPCR assays for norovirus could have a significantly reduced genome amplification efficiency than the other assays depending on viral sequences present in clinical and environmental samples (**Fig. 2** and **3**). Therefore, virus surveillance should be conducted with an assay that can cover a wide range of viral sequences that may be introduced to a community. We found that the probability of detection determined *in silico* with the up-to-date viral sequences could be used to evaluate the likelihood of reduced quantification efficiency for clinical testing or wastewater surveillance.

The calculated probability of detection may not be the perfect parameter to describe the PCR amplification efficiency because it assumes that the perfect match between a PCR assay and a virus genome is a prerequisite for the amplification. In fact, the PCR assay could still detect viruses even with mismatches between the genome and primers. The probability of detection does not differentiate the varying impacts of mismatches on the amplification of the target sequence. It is currently challenging to quantitatively evaluate mismatch because the number, location, and type of mismatch or their combinations have complex impacts on PCR amplification efficiency (57–59). Despite the limitation, the probability of detection would still be helpful in evaluating nucleic acid-based assays because it is better to minimize the number of mismatches. For example, the viral sequences from the databases (e.g., Genbank or GISAID) are probably not perfectly representative of the true viral diversity in reality, meaning that there may be a viral sequence with mismatches on annealing sites that have not been reported in the database yet. Indeed, the Genbank database did not include sequences that are identical to 12 norovirus sequences of our clinical samples (out of 15 sequences). We sequenced norovirus RNA in the clinical samples (from 229 to 833 bases) and the most similar sequences in Genbank showed from one to ten mismatches (**Table S5**). In addition, mutations frequently occur in viruses, which could eventually lead to the appearance of new mismatches on the annealing sites (60). The unexpected extra mismatch could result in failure of PCR analysis (i.e., false negative).

The current practice for reporting PCR assay results, such as Minimum Information for Publication of Quantitative Real-Time PCR Experiments (MIQE) guidelines, focuses on the quality control and reproducibility of data and does not demand thorough consideration of the impact of genome diversity on RT-qPCR analysis. For example, the MIQE list requests to present target gene information, but a great level of diversity is also found at a gene level too, as shown in segmented genes of RV and IV (**Fig. S5**). This means the target gene may not be enough information to address genome diversity. In this study, we propose that the probability of detection be used to evaluate the performance of nucleic acid-based assays for virus surveillance in advance. The probability of detection of nucleic acid-based assay can be simply calculated *in silico*, and we published a code that calculates the probability of detection of degenerate RT-qPCR (github.com/Nguyen205/In-silico-analysis-for-degenerate-qPCR-assay). This tool will enable people to easily evaluate their nucleic acid-based assays and improve the reliability of virus surveillance.

## Data Availability

All data produced in the present study are available upon reasonable request to the authors

## Acknowledgment

We acknowledge the funding from the Grainger College of Engineering, the JUMP-ARCHES program of OSF Healthcare in conjunction with the University of Illinois, and the VinUni Illinois Smart Health Center. We thank Brad Bennett and Bruce Rabe at the Urbana-Champaign Sanitary District and Haley Turner and Travis Ramme at the Rantoul Wastewater Treatment Plant for providing us with influent wastewater. The authors also acknowledge Kip Stevenson for sampling deployment, and Yuqing Mao, Matthew Robert Loula, Aashna Patra, Kristin Joy Anderson, Mikayla Diedrick, Hubert Lyu, Hamza Elmahi Mohamed, Jad R Karajeh, Runsen Ning, Rui Fu, and Kyukyoung Kim for sewage sampling and processing. We also acknowledge Dr. Awais Vaid for guidance on sampling site selection, and Dr. Geltz at the Illinois Department of Public Health for providing us with fecal samples.

