## Supplemental Figures and Tables for "Improved performance of nucleic acid-based assays for genetically diverse norovirus surveillance"

**Two text, five figures, and thirteen tables are included.**

### Viral sequence download for *in silico* analysis

Norovirus sequences in a fasta format (n=637) were downloaded from the nucleotide database of Genbank with a keyword of "human norovirus" and a filter of sequence length longer than 5000 bases. Rotavirus A species' complete gene sequences (11 segmented genes and n=18327 in total) were downloaded from Genbank with a keyword of "human rotavirus A". Respiratory syncytial virus (RSV) complete sequences (n=1798) were downloaded from Genbank with a keyword "human respiratory syncytial virus USA" and a filter of sequence length between 10000 and 20000 bases. Influenza A virus (IAV) segmented sequences were obtained from the influenza virus database of Genbank with searching conditions of nucleotide, influenza A type, human as a host, USA as a country, full-length sequence, and collection date between 2019 and 2022. The number of each segmented RNA ranged from 10953 to 11880. Adenovirus type 41 complete sequences (n=126) were acquired from the nucleotide database of Genbank with a keyword of "human adenovirus type 41" and a filter of sequence length between 30000 and 40000 bases. SARS-CoV-1 complete sequences (n=151) were downloaded from the Genbank with a keyword of "severe acute respiratory syndrome-related coronavirus" and a filter of sequence length between 29000 and 31000 and release date from 2002 to 2006. MERS complete sequences (n=720) were obtained from Genbank too with a keyword of "Middle East respiratory syndrome-related coronavirus" and a filter of sequence length between 29000 to 31000. Ebola virus complete sequences (n=1859) were acquired from Genbank with a keyword of "Ebola virus" and a filter of sequence length between 18000 and 20000 and a release date from 2013 to 2016. SARS-CoV-2 complete sequences (n=356) were obtained from the EpiCoV database of GISAID that were reported from Champaign county (Illinois, USA) with two filters of "complete" and "high coverage". Mpox complete sequences (n=41) were downloaded from EpiPox database of GISAID that were deposited from Illinois, USA with two filters of "complete" and "high coverage".

### Alignment for viral species with a long sequence length and/or numerous sequences

Specifically, complete RSV genomes (about 15,000 bases) were cut into five pieces, each of which was less than 4,000 bases long, complete AdV genomes (about 35,000 bases) were divided into four fragments (<10,000 bases), complete SARS-CoV-1 sequences (about 30,000 bases) were sliced into three fragments (<12,000 bases), complete MERS sequences (about 30,000 bases) were fragmented to eight pieces (<5,000 bases), complete SARS-CoV-2 genomes (about 30,000 bases) were fragmented into six pieces (< 6,000 bases), complete MERS sequences (about 30000 bases) were taken apart into eight pieces (<5000), Ebola virus sequences (about 19,000 bases) were cut into six pieces (<5,000), and complete Mpox genomes (about 200,000 bases) were sliced into 50 pieces (<5,000 bases). The alignments from each piece of the genome were then combined back to obtain the complete alignment with about 1,000 bases being overlapped by two adjacent fragments.

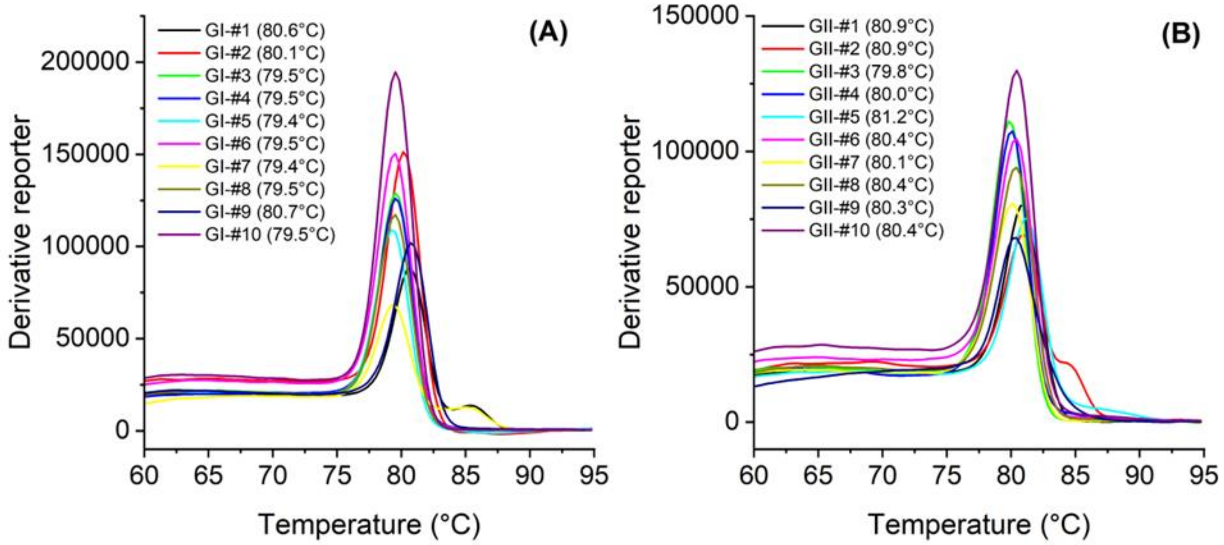


**Fig. S1.** Melting curves of SYBR-based RT-qPCR assays: (A) A1 assay for GI genogroup and (B) B1 assay for GII genogroup.

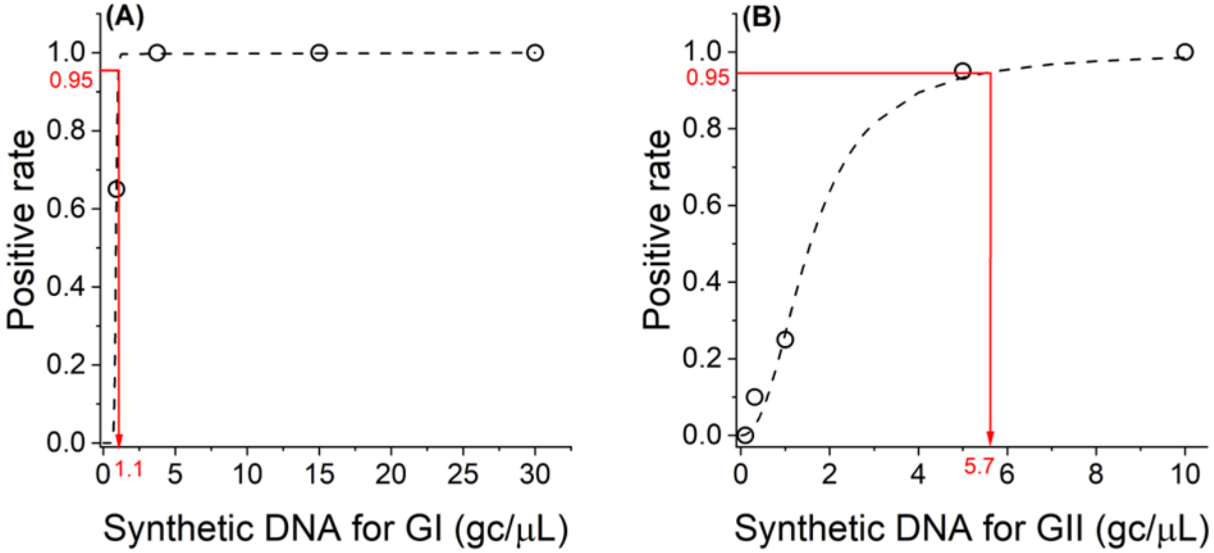


**Fig. S2.** Limit of detection (LOD) of A1 assay **(A)** and B1 assay **(B)**. Serial dilutions of synthetic DNA for GI and GII were used for the A1 and B1 assay, respectively. The LODs were the DNA concentration at which the positive rate was 0.95 (red solid lines).

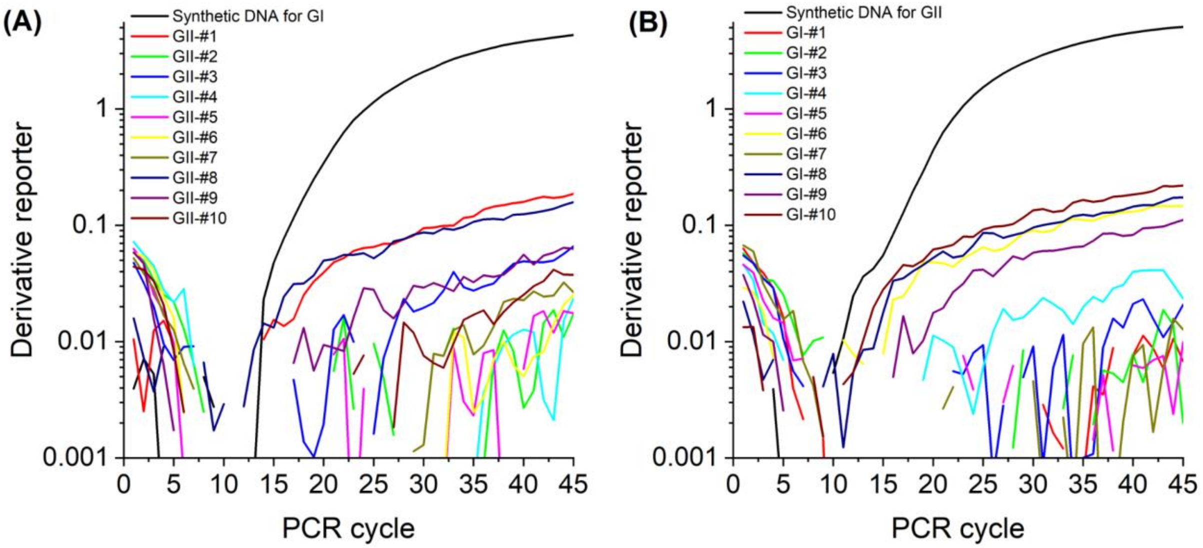


**Fig. S3.** Specificity test of RT-qPCR assays for (A) GI and (B) GII genogroup. (A) The A1 assay for GI genogroup was applied to synthetic DNA control for GI genogroup and ten clinical samples of GII genogroup. (B) The B1 assay for GII genogroup was applied to synthetic DNA control for GII genogroup and ten clinical samples of GI genogroup.

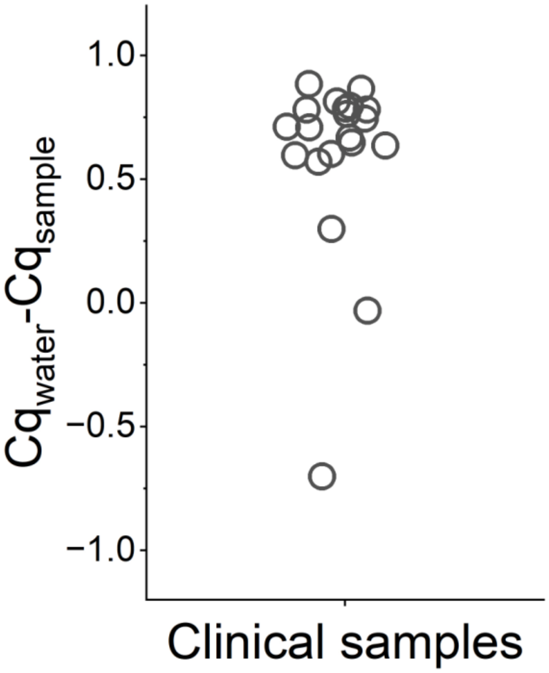


**Fig. S4**. Inhibition test for clinical samples of norovirus GI and GII genogroups (n=20).

**Table S1.** Detailed information about RT-qPCR assays for norovirus GI and GII genogroups

| Target | Assay | type | Sequence (5' to 3') | Degeneracy | Location | Amplicon size (bp) | Length | GC (%) | Tm (ºC)[1)] | Annealing temperature (ºC) | Probability of detection |
|---|---|---|---|---|---|---|---|---|---|---|---|
| GI[11)] | A1[7)] | For | GCHATRTTYCGYTGGATG | 24 | 5283-5300[2)] | 93 | 18 | 49.1 | Min: 53.6<br>Mean: 57.9<br>Max: 63.6 | 53 | 48/48 (100%) |
| | | Probe | TGGACAGGRGAYCGCRATCT | 8 | 5322-5341 | | 20 | 57.5 | Min: 63.5<br>Mean: 66.1<br>Max: 69.7 | | |
| | | Rev | TTAGACGCCATCATCATT | 1 | 5358-5375 | | 18 | 38.9 | 56.2 | | |
| | A2[8)] | For | GCYATGTTCCGYTGGATGC | 4 | 5283-5301[2)] | 97 | 19 | 57.9 | Min: 61.5<br>Mean: 63.4<br>Max: 66.4 | 57 | 27/48 (56%) |
| | | Probe | TCGGGCAGGAGATYGCGRTCYC | 8 | 5329-5350 | | 22 | 65.9 | Min: 68.1<br>Mean: 70.4<br>Max: 73.5 | | |
| | | Rev | GTCCTTAGACGCCATCATCATT | 1 | 5358-5379 | | 22 | 45.5 | 62.3 | | |
| | A3[9)] | For | GGAGATCGCRATCTCCTGCC | 2 | 5312-5331[4)] | 103 | 20 | 62.5 | Min: 64.8<br>Mean: 65.1<br>Max: 66.4 | 60 | 4/48 (8%) |
| | | Probe | GGGGCGTCCTTAGACGCCATCATCATTTA | 1 | 5335-5363 | | 29 | 52.7 | 70.6 | | |
| | | Rev | CTCYGGTACCAGCTGGCC | 2 | 5397-5414 | | 18 | 69.4 | Min: 64.2<br>Mean: 64.9<br>Max: 66.7 | | |
| GII[12)] | B1[7)] | For | TTYAGRTGGATGAGRTTYTC | 16 | 5012-5030[6)] | 86 | 19 | 43.9 | Min: 52.7<br>Ave: 55.8<br>Max: 60.3 | 53 | 542/561 (97%) |
| | | Probe | ACDTGGGAGGGYGATCRCAAT | 12 | 5042-5061 | | 20 | 55 | Min: 63.3<br>Ave: 65.3<br>Max: 68.3 | | |
| | | Rev | YMGAYGCCATCHTCATTC | 24 | 5080-5097 | | 18 | 49.1 | Min: 56.3<br>Ave: 57.5<br>Max: 60.5 | | |
| | B2[10)] | For | ATGTTCAGRTGGATGAGRTTCTCWGA | 8 | 5009-5034[6)] | 89 | 26 | 42.3 | Min: 64.2<br>Mean: 65.3<br>Max: 67.4 | 60 | 174/561 (31%) |
| | | Probe | AGCACGTGGGAGGGCGATCG | 1 | 5039-5058 | | 20 | 70.0 | 70.3 | | |
| | | Rev | TCGACGCCATCTTCATTCACA | 1 | 5077-5097 | | 21 | 47.6 | 64.1 | | |
| | B3[9)] | For | GTGGGATGGACTTTTACGTGCCAA | 1 | 4974-4997[6)] | 129 | 24 | 50.0 | 66.9 | 60 | 91/561 (16%) |
| | | Probe | AGCCAGATTGCGATCGCCCTCC | 1 | 5051-5072 | | 22 | 63.6 | 70.2 | | |
| | | Rev | CGTCAYTCGACGCCATCTTCATTCA | 2 | 5078-5102 | | 25 | 50.0 | Min: 67.3<br>Mean: 67.4<br>Max: 68.4 | | |

1) Melting temperatures were calculated with the qPCR parameter sets of OligoAnalyzer (Integrated DNA Technology), which consider 0.2 μM of oligo concentration, 50 mM of $Na^+$ concentration, 3 mM of $Mg^{2+}$ concentration, and 0.8 mM of dNTPs concentration.
2) Reference sequence (Genbank ID: KX396056.1)
3) Reference sequence (Genbank ID: MW305482.1)
4) Reference sequence (Genbank ID: MW243609.1)
5) Reference sequence (Genbank ID: MW305489.1)
6) Reference sequence (Genbank ID: MW661246.1)
7) Developed in this study
8) Adopted from Wolf et al. (2010)(1)
9) Adopted from Liu et al. (2020)(2)
10) Adopted from Loisy et al. (2005)(3)
11) The sequence of a standard sample for the ORF1 and ORF2 genes of norovirus GI genogroup (Integrated DNA Technologies, USA): 5’-CAGAAAAATCTCCAGTAAAGTTATACAGGAGATAAGGACCGGTGGCTTAGAAATGTATGTACCAGGTTGGCAGGCCATGTTCCGCTGGATGCGCTTCCATGATCTCGGATTGTGGACAGGAGATCGCAATCTCCTGCCCGAATTCGTAAATGATGATGGCGTCTAAGGACGCTACACCAAGCGCAGATGGCGCCACTGGCGCCGGCCAGCTGGTACCGGAGGTTAATACAGCTGACCCTATACCCATTGACCCTGTGGCTGGCTCCTCTACAGCCCTTGCCACTGCGGGCCAAGTTAATT-3’ (GenBank accession number: MW305499.1)
12) The sequence of a standard sample for the ORF1 and ORF2 genes of norovirus GII genogroup (Integrated DNA Technologies, USA): 5’-ACCCCACTCTCAAAGACCCATACAGCTCATGGCACTGCTTGGTGAGGCCTCCCTTCACGGACCCTCTTTCTACAGCAAAATCAGTAAATTGGTCATAACTGAACTCAAAGAAGGTGGGATGGACTTTTACGTGCCAAGGCAGGAACCCATGTTCAGGTGGATGAGGTTCTCTGACTTGAGCACGTGGGAGGGCGATCGCAATCTGGCTCCCAGTTTTGTGAATGAAGATGGCGTCGAATGACGCCGCTCCATCTACTGATGGTGCAGCCGGCCTCGTGCCAGAAAGTAACAGTGAGGTCA-3’ (GenBank accession number: MW661246.1)

**Table S2.** The checklist from MIQE guidelines and relevant information for this study

| Item to check | Location |
|---|---|
| 1. Experimental design | |
| Definition of experimental and control groups | Materials and Methods |
| Number within each group | Materials and Methods; Each figure |
| 2. Sample | |
| Description | Clinical sample collection and processing<br>Sewage sample collection and processing |
| Volume/mass of sample processed | Clinical sample collection and processing<br>Sewage sample collection and processing |
| Processing procedure | Clinical sample collection and processing<br>Sewage sample collection and processing |
| Sample storage conditions and duration | Clinical sample collection and processing<br>Sewage sample collection and processing |
| 3. Nucleic acid extraction | |
| Procedure and/or instrumentation | Clinical sample collection and processing<br>Sewage sample collection and processing |
| Name of kit and details of any modifications | Clinical sample collection and processing<br>Sewage sample collection and processing |
| Contamination assessment (DNA or RNA) | RT-qPCR protocol for norovirus quantification |
| Nucleic acid quantification | RT-qPCR protocol for norovirus quantification |
| Instrument and method | Clinical sample collection and processing<br>Sewage sample collection and processing |
| Inhibition testing ($C_q$ dilutions, spike, or other) | Clinical sample collection and processing<br>Sewage sample collection and processing<br>Fig. S4 and Oh et al. (2022)(4) |
| 4. Reverse transcription | |
| Complete reaction conditions | RT-qPCR protocol for norovirus quantification, Table S1 |
| Amount of RNA and reaction volume | RT-qPCR protocol for norovirus quantification, Table S1 |
| Reverse transcriptase and concentration | RT-qPCR protocol for norovirus quantification, Table S1 |
| Temperature and time | RT-qPCR protocol for norovirus quantification, Table S1 |
| Manufacturer of reagents and catalogue numbers | RT-qPCR protocol for norovirus quantification, Table S1 |
| 5. qPCR target information | |
| Gene symbol | RT-qPCR protocol for norovirus quantification, Table S1 |

| | |
|---|---|
| Sequence accession number | RT-qPCR protocol for norovirus quantification, Table S1 |
| Location of amplicon | RT-qPCR protocol for norovirus quantification, Table S1 |
| Amplicon length | RT-qPCR protocol for norovirus quantification, Table S1 |
| In silico specificity screen (BLAST, and so on) | RT-qPCR protocol for norovirus quantification, Table S1 |
| 6. qPCR oligonucleotides | |
| Primer sequences | RT-qPCR protocol for norovirus quantification, Table S1 |
| Manufacturer of oligonucleotides | RT-qPCR protocol for norovirus quantification, Table S1 |
| 7. qPCR protocol | |
| Complete reaction conditions | RT-qPCR protocol for norovirus quantification, Table S1 |
| Reaction volume and amount of DNA | RT-qPCR protocol for norovirus quantification, Table S1 |
| Primer (probe) concentrations | RT-qPCR protocol for norovirus quantification, Table S1 |
| Polymerase identity and concentration | RT-qPCR protocol for norovirus quantification, Table S1 |
| Buffer/kit identity and manufacturer | RT-qPCR protocol for norovirus quantification, Table S1 |
| Manufacturer of plates/tubes and catalog number | RT-qPCR protocol for norovirus quantification, Table S1 |
| Complete thermocycling parameters | RT-qPCR protocol for norovirus quantification, Table S1 |
| Manufacturer of qPCR instrument | RT-qPCR protocol for norovirus quantification, Table S1 |
| 8. qPCR validation | |
| Specificity (gel, sequence, melt, or digest) | Degenerate RT-qPCR assay design for genetically variable norovirus surveillance |
| For SYBR Green I, $C_q$ of the NTC | RT-qPCR protocol for norovirus quantification |
| Calibration curves with slope and *y* intercept | RT-qPCR protocol for norovirus quantification |
| PCR efficiency calculated from slope | RT-qPCR protocol for norovirus quantification |
| *r*2 of calibration curve | RT-qPCR protocol for norovirus quantification |
| Linear dynamic range | RT-qPCR protocol for norovirus quantification |
| $C_q$ variation at LOD | Degenerate RT-qPCR assay design for genetically variable norovirus surveillance<br>RT-qPCR protocol for norovirus quantification<br>Fig. S2 |
| Evidence for LOD | Degenerate RT-qPCR assay design for genetically variable norovirus surveillance<br>RT-qPCR protocol for norovirus quantification<br>Fig. S2 |
| 9. Data analysis | |
| qPCR analysis program (source, version) | RT-qPCR protocol for norovirus quantification |

| | |
|---|---|
| Method of $C_q$ determination | RT-qPCR protocol for norovirus quantification |
| Outlier identification and disposition | RT-qPCR protocol for norovirus quantification |
| Results for NTCs | RT-qPCR protocol for norovirus quantification |
| Description of normalization method | RT-qPCR protocol for norovirus quantification |
| Number and concordance of biological replicates | RT-qPCR protocol for norovirus quantification; each Figure |
| Number and stage of technical replicates | RT-qPCR protocol for norovirus quantification |
| Repeatability (intraassay variation) | RT-qPCR protocol for norovirus quantification |
| Statistical methods for results significance | Statistical analysis |
| Software (source, version) | RT-qPCR protocol for norovirus quantification |
| Data transparency | Raw data available upon request |

**Table S3.** Summary of stool specimen positive for norovirus

| Sample ID | Genotype[1)] | Primers | Collection date | Collection location |
|---|---|---|---|---|
| GI-#1 | - | - | 12/18/2018 | Champaign |
| GI-#2 | GI.3 | GI_Set3 | 7/16/2019 | McHenry |
| GI-#3 | GI.3 | GI_Set3 | 8/27/2019 | DuPage |
| GI-#4 | GI.7 | GI_Set3 | 10/25/2019 | McHenry |
| GI-#5 | GI.7 | GI_Set3 | 10/27/2019 | McHenry |
| GI-#6 | GI.7 | GI_Set3 | 10/27/2019 | McHenry |
| GI-#7 | - | - | 10/27/2019 | McHenry |
| GI-#8 | GI.7 | GI_Set3 | 10/26/2019 | McHenry |
| GI-#9 | - | - | 2/19/2022 | McHenry |
| GI-#10 | GI.3 | GI_Set3 | 6/15/2022 | Pike |
| GII-#1 | GII.8 | GII_Set9 | 8/27/2019 | DuPage |
| GII-#2 | - | - | 9/5/2019 | DuPage |
| GII-#3 | GII.17 | GII_Set3 | 11/1/2019 | McHenry |
| GII-#4 | GII.17 | GII_Set3 | 11/2/2019 | McHenry |
| GII-#5 | GII.4 | GII_Set3 | 1/30/2020 | McHenry |
| GII-#6 | GII.4 | GII_Set11 | 2/11/2020 | McHenry |
| GII-#7 | GII.4 | GII_Set11 | 2/26/2021 | Sangamon |
| GII-#8 | GII.4 | GII_Set11 | 2/25/2021 | Sangamon |
| GII-#9 | - | - | 3/3/2021 | Sangamon |
| GII-#10 | GII.4 | GII_Set3 | 3/4/2021 | Sangamon |

1) Norovirus sequences determined by Sanger sequencing were input the Norovirus genotyping tool to determine the genotypes of clinical samples.

**Sanger sequencing**

Clinical samples were analyzed by Sanger sequencing to obtain viral sequences. First, we synthesized complementary DNA (cDNA) from the norovirus genomic RNA using the First Strand cDNA Synthesis Kit (New England BioLabs, USA). Six microliters of RNA samples were mixed with 10 µL of M-MuLV Reaction Mix, 2 µL of M-MuLV Enzyme Mix, and 2 µL of 10 µM of reverse primers. The mixture was incubated at 42℃ for 60 minutes for cDNA synthesis, followed by 80℃ for 10 minutes for enzyme inactivation. The 3.5 mL of cDNA was then mixed with 0.5 µL of Phusion DNA polymerase, 10 µL of 5X Phusion HF buffer, 2.5 µL of 10 µM forward primer, 2.5 µL of 10 µM reverse primer, 1 µL of 10 mM dNTPs, 30 µL of nuclease-free water (Phusion® High-Fidelity PCR Kit, MA, USA). This 50 µL of PCR cocktail was incubated at 98°C for 30 seconds for initial denaturation, 40 cycles of denaturation at 98°C for 10 seconds, annealing at various temperatures for each primer set (**Table S4**) for 30 seconds, extension at 72°C for 30 seconds, and 72°C for 10 minutes (final extension). The PCR amplicon was purified using QIAquick PCR Purification Kit (QIAGEN, Germany) following the manufacturer's protocol, and the PCR amplicon was eluted in 30 µL of nuclease-free water. In addition, the PCR amplicon was further cleaned up by ExoSAP-IT™ Express PCR Product Cleanup Reagent (Applied Biosystems, MA, USA) following the manufacturer's procedure. The double-stranded DNA (dsDNA) concentration of the amplicon was determined by Qubit 2.0 fluorometer (Invitrogen, USA). The Core DNA Sequencing Facility at the University of Illinois Urbana-Champaign analyzed the samples through Sanger sequencing. The norovirus genome sequences were finalized after examining the sequencing chromatogram (i.e., dye terminator peaks, the baseline, and the sequence text) with FinchTV (version 1.4.0).

**Table S4**. Summary of primers for sanger sequencing

| Sample ID | Primer type | Sequence (5' to 3') | Length (bp) | Tm (ºC)[1] | GC (%) | Amplicon location (size) |
|---|---|---|---|---|---|---|
| GI_Set3[2] | Forward primer | TCATTTTATGGTGATGATGAAAT | 23 | 48.5 | 26.1 | 4889-5550 (662 bp) |
| | Reverse primer | AGGGGTCAATCATATTAACTTG | 22 | 50.1 | 36.4 | |
| GI_Set9[3] | Forward primer | CCTTGCACATCTCAGGTGAATA | 22 | 54.7 | 45.5 | 4703-5572 (870 bp) |
| | Reverse primer | TGAGGCCCTAACTGCAAATC | 20 | 55.1 | 50.0 | |
| GII_Set3[4] | Forward primer | TTCTATGGTGATGATGAGATTGT | 23 | 51.5 | 34.8 | 4612-5243 (632 bp) |
| | Reverse primer | CTAATCCAGGGGTCAATTACAT | 22 | 52.0 | 40.9 | |
| GII_Set9[5] | Forward primer | CAATAGCACACTGGATCCTAAC | 22 | 53.1 | 45.5 | 4459-5324 (866 bp) |
| | Reverse primer | CTAGCCAGATGTGCAAGATAAG | 22 | 53.1 | 45.5 | |
| GII_Set11[6] | Forward primer | CCCATTCTCAAAGACCCATACA | 22 | 54.5 | 45.5 | 4865-5693 (829 bp) |
| | Reverse primer | TGAGAACTCGGCACGAAAC | 19 | 55.3 | 52.6 | |
| GII_Set14[7] | Forward primer | GATTTGAATGGTCTCACATTCTTG | 24 | 52.4 | 37.5 | 4744-5336 (593 bp) |
| | Reverse primer | TTCTGGACCTAACTCTAAATCTAAC | 25 | 51.8 | 36.0 | |

1) Melting temperatures were calculated with the default parameter sets of OligoAnalyzer (Integrated DNA Technology), which consider 0.25 μM of oligo concentration and 50 mM of $Na^+$ concentration.
2) Reference sequence (Genbank ID: MT031988.1)
3) Reference sequence (Genbank ID: MW305499.1)
4) Reference sequence (Genbank ID: OP727614.1)
5) Reference sequence (Genbank ID: MW305576.1)
6) Reference sequence (Genbank ID: MZ478141.1)
7) Reference sequence (Genbank ID: OP686904.1)

**Table S5.** Norovirus sequences of clinical samples

| Sample # | Confirmed viral sequences[1)] | Note[2)] |
|---|---|---|
| GI-1 | - | - |
| GI-2 | 5’-<br>CAACAGATATAGAATTTGACCCAATCAAACTGACACAAATACTGAAGGAATATGGTTTGAAACCCACAA<br>GACCTGACAAAACTGATGGCCCAATTATAGTCAGACAGCAAGTGGATGGCCTGGTCTTCCTCCGGCGCA<br>CCATCTCTAAGGATGCTATTGGATACCAGGGACGGCTCGATCGCAATTCCATTGAAAGACAGCTATGGTG<br>GACTCGCGGGCCAAATCACGATGACCCGTTTGAGACACTGGTCCCGCATTCACAGAGGAAGGTCCAATT<br>AGTATCTCTGCTTGGTGAAGCAGCACTTCATGGTGAAAAGTTCTACAGAAAGATAGCCGGCAGAGTTATT<br>CAAGAAGTCAAAGAGGGGGGGCTTGAAATCTACATTCCCGGCAGGCAGGCCATGTTCCGCTGGATGCGC<br>TTTCATGATCTGAGTTTGTGGACAGGGGACCGCGATCTCCTGCCCGATTATGTAAATGATGATGGCGTCT<br>AAGGACGCCCCAACAAACATGGATGGCACCAGTGGTGCCGGCCAGCTGGTACCAGAGGCAAATACAGC<br>TGAGCCTATATCAATGGAGCCTGTGGCTGGGGCAGCGACAGCTGCCGCAACCGCTGGCCAAGTTAAT-3’ | 616 out of 621 (99%) nucleotides match to MT031988.1 |
| GI-3 | 5’-<br>CCGATCATTGTAAGGCAACAGGTTGATGGCTTGGTTTTCCTCCGGCGCACCATCTCAAAGGATGCCATCG<br>GGTACCAGGGCCGGCTTGACCGTAATTCCATTGAAAGACAGCTCTGGTGGACCCGGGGGCCCAACCATG<br>ATGATCCATTTGAAACCTTGGTCCCACACCCGCAGAGGAAGGTCCAACTGATATCCCTGCTGGGTGAAGC<br>TGCACTCCATGGTGAGAAGTTCTACAGAAAGATAGCCAGTAGGGTGATCCAGGAGGTTAAAGAAGGAG<br>GATTGGAAATTTACATCCCTGGGTGGCAGGCCATGTTCCGCTGGATGCGATTCCATGATTTGAGCTTGTG<br>GACAGGAGACCGCGATCTCTTGCCCGATTATGTAAATGATGATGGCGTCTAAGGACGCCCCAACAAACA<br>TGGATGGCACCAGTGGTGCCGGTCAGCTGGTACCAGAGGCAAATACAGCTGAACCTATATCAATGGATC<br>CAGTAGCTGGAGCCGCAACAGCGGTTGCAACTG-3’ | 518 out of 518 (100%) nucleotides match to MN922735.1 |
| GI-4 | 5’-<br>AACCATGATGATCCCTTTGAGACATTAATACCCCATCAACAAAGAAAGATTCAATTGATTTCCTTACTTG<br>GTGAGGCTGCGCTCCACGGAGAGAAATTCTATAGAAAGATTGCCAACAGAGTCATACAGGAAGTCAAAG<br>AAGGGGGCCTTGAGCTCTATATACCAGGTTGGCAGGCCATATTCCGCTGGATGCGTTTCCATGACTTGAG<br>CTTGTGGACAGGAGATCGCAATCTCCTGCCCGATTATGTAAATGATGATGGCGTCTAAGGACGCCCCCTC<br>AAACATGGATGGCACTAGTGGTGCCGGTCAGCTGGTTCCAGAGGTTAATGCAGCTGAACCCCTACCCCTT<br>GAGCCGGTGGTGGGTGCCGCAACTGCGGTGGCCACTGCTGGGCAAGTTAA-3’ | 397 out of 398 (99%) nucleotides match to LC646334.1 |
| GI-5 | 5’-<br>CGCAACTCCATTGAAAGACAATTATGGTGGACCCGGGGCCCAAACCATGATGATCCCTTTGAGACATTA<br>ATACCCCATCAACAAAGAAAGATTCAATTGATTTCCTTACTTGGTGAGGCTGCGCTCCACGGAGAGAAAT<br>TCTATAGAAAGATTGCCAACAGAGTCATACAGGAAGTCAAAGAAGGGGGCCTTGAGCTCTATATACCAG<br>GTTGGCAGGCCATATTCCGCTGGATGCGTTTCCATGACTTGAGCTTGTGGACAGGAGATCGCAATCTCCT<br>GCCCGATTATGTAAATGATGATGGCGTCTAAGGACGCCCCCTCAAACATGGATGGCACTAGTGGTGCCG<br>GTCAGCTGGTTCCAGAGGTTAATGCAGCTGAACCCCTACCCCTTGAGCCGGTGGTGGGTGCCGCAACTGC<br>GGTGGCCACTGCTGGGCAAGTTAAT-3’ | 441 out of 442 (99%) nucleotides match to LC646334.1 |
| GI-6 | 5’-<br>CAGGCCATATTCCGCTGGATGCGTTTCCATGACTTGAGCTTGTGGACAGGAGATCGCAATCTCCTGCCCG<br>ATTATGTAAATGATGATGGCGTCTAAGGACGCCCCCTCAAACATGGATGGCACTAGTGGTGCCGGTCAG<br>CTGGTTCCAGAGGTTAATGCAGCTGAACCCCTACCCCTTGAGCCGGTGGTGGGTGCCGCAACTGCGGTGG<br>CCACTGCTGGGCAAGTTAAT-3’ | 229 out of 229 (100%) nucleotides match to MN421785.1 |
| GI-7 | - | - |
| GI-8 | 5’-<br>GAAGCATCAAATAGATGGGTTAGTTTTTCTGAGGCGCACTATATCAAAAGATGCTGCTGGCTACCAAGG<br>GCGCTTGGACCGCAACTCCATTGAAAGACAATTATGGTGGACCCGGGGCCCAAACCATGATGATCCCTTT<br>GAGACATTAATACCCCATCAACAAAGAAAGATTCAATTGATTTCCTTACTTGGTGAGGCTGCGCTCCACG<br>GAGAGAAATTCTATAGAAAGATTGCCAACAGAGTCATACAGGAAGTCAAAGAAGGGGGCCTTGAGCTCT<br>ATATACCAGGTTGGCAGGCCATATTCCGCTGGATGCGTTTCCATGACTTGAGCTTGTGGACAGGAGATCG<br>CAATCTCCTGCCCGATTATGTAAATGATGATGGCGTCTAAGGACGCCCCCTCAAACATGGATGGCACTAG<br>TGGTGCCGGTCAGCTGGTTCCAGAGGTTAATGCAGCTGAACCCCTACCCCTTGAGCCGGTGGTGGGTGCC<br>GCAACTGCGGTGGCCACTGCTG-3’ | 509 out of 510 (99%) nucleotides match to LC646334.1 |
| GI-9 | - | |
| GI-10 | 5’-<br>TAGTCTCAACAGATATTGAATTTGACCCAAACAGGTTAACACAAGTTCTAAGAGAGTATGGCTTAAAGC<br>CCACAAGACCTGACAAGACTGATGGCCCAATCATTGTGAGACAGCAAGTTGATGGCTTGGTTTTCCTCCG<br>GCGCACCATTTCGAAAGATGCCATTGGATACCAGGGACGCCTCGACCGAAATTCCATTGAGAGACAGCT<br>CTGGTGGACTCGTGGGCCCAACCATGATGATCCATTTGAAACCTTAGTCCCACATACACAGAGAAAGGTT<br>CAGCTAATATCCCTACTAGGTGAAGCTGCACTCCATGGTGAGAAATTCTACAGAAAGATAGCCAGTAGG<br>GTGATCCAGGAAGTCAAAGAGGGGGGGTTGGAAGTTTACATCCCTGGGTGGCAGGCCATGTTCCGCTGG<br>ATGCGATTCCATGATTTGAGCTTGTGGACAGGAGACCGCGATCTCTTGCCCGATTATGTAAATGATGATG<br>GCGTCTAAGGACGCCCCAACAAACATGGATGGCACCAGTGGTGCCGGTCAGCTGGTACCAGAGGCGAAT<br>ACAGCTGAACCTATATCAATGGATCCAGTGGCTGGAGCCGCAACAGCGGTCGCTACTGCTGGACAAGTT<br>AATA-3’ | 620 out of 628 (99%) nucleotides match to MN922741.1 |

| | | |
|---|---|---|
| GII-1 | 5'-<br>CTCTTAGTGCTATGTCTGAGGTCTCTGGTCTTTCCCCTGAGGTTGTGCAAGCCAACTCCTGTTTCTCATTCT<br>ATGGGGATGATGAAATAGTCAGCACAGATATAAACCTAGACCCAGAAAAACTCACCAGGAAACTGAGG<br>GAGTATGGCCTCGTCCCAACAAGGCCAGACAAAACTGAGGGCCCACTTGTGATCACTCAGGATTTGAAT<br>GGTCTCACATTCTTGAGGCGAACCATAGTGCGGGACCCCGCAGGTTGGTTTGGAAAATTGGATCGTGATT<br>CCATTCTAAGGCAGTTATACTGGACCAGAGGACCCAATCATGAGAACCCCTTTGAAAGTATGATTCCCCA<br>CTCCCAGAGAGCAACCCAGTTAATGGCCCTTCTTGGGGAAGCCTCGTTGCATGGTCCCCAATTTTACAAG<br>AAGGTGAGTAAAATGGTCATCAGTGAGATCAAGAGTGGTGGTCTGGAGTTTTACGTGCCCAGACAGGAG<br>GCCATGTTTAGATGGATGAGATTTTCAGACCTCAGCACGTGGGAGGGCGATCGCAATCTGGCTCCCGAG<br>AATGTGAATGAAGATGGCGTCGAATGACGCAGCTCCATCGAATGATGGCGCGGCTGGCCTCGTACCAGA<br>GATCAACCATGAGGTCATGGCCATAGAGCCTGTTGCAGGGGCCTCTCTAGCAGCCCCTGTCGTAGGACA<br>ACTTAATATAATTGATCCCTGGATTAGAAATAATTTTGTACAAGCCCCTGCTGGAGAATTCACTGTTTCG<br>CCTAGAAATGCTCCAGGTGAATTTTTGTTAGATTTAGAGTTAGGTCCAGAATTGAATCCTTATCTTGCA-3' | 831 out of 833 (99%) nucleotides match to OP686904.1 |
| GII-2 | - | - |
| GII-3 | 5'-<br>TCCTCCGCCGAACAGTCACCCGTGATCCAGCAGGTTGGTTTGGAAAGTTGGACCAAAACTCCATCCTCAG<br>GCAGTTGTACTGGACAAGAGGACCCAACCATGAAGACCCCAGTGAGACCATGATACCACACGCACAAA<br>GACCTGTGCAGCTCATGGCACTACTAGGAGAATCCTCCCTACATGGACCCTCATTTTACAGCAAGGTTAG<br>CAAATTAGTCATATCTGAACTTAAAGAGGGAGGAATGGATTTTTATGTGCCCAGACAAGAGTCAATGTTC<br>AGGTGGATGAGGTTCTCAGATCTAAGCACATGGGAGGGCGATCGCAATCTGGCTCCCAGTTTTGTGAAT<br>GAAGATGGCGTCGAATGACGCCGCTCCATCTAATGATGGTGCTGCTGGTCTCGTACCAGAGGGCAACAA<br>CGAGACCCTTCCCCTAGAACCAGTTGCGGGCGCAGCTATAGCCGCACCCGTCACTGGCCAAAATAA-3' | 482 out of 482 (100%) nucleotides match to KT326180.1 |
| GII-4 | 5'-<br>TGATGATGAGATTGTGAGCACAGACATAAAATTGGACCCAGAAAAATTGACCGCAAAGCTCAAAGAATA<br>TGGCCTTAAACCCACTCGGCCCGACAAAACTGAGGGGCCGTTGGTGATTAGTGAGGACCTGAATGGGTT<br>GACTTTCCTCCGCCGAACAGTCACCCGTGATCCAGCAGGTTGGTTTGGAAAGTTGGACCAAAACTCCATC<br>CTCAGGCAGTTGTACTGGACAAGAGGACCCAACCATGAAGACCCCAGTGAGACCATGATACCACACGCA<br>CAAAGACCTGTGCAGCTCATGGCACTACTAGGAGAATCCTCCCTACATGGACCCTCATTTTACAGCAAGG<br>TTAGCAAATTAGTCATATCTGAACTTAAAGAGGGAGGAATGGATTTTTATGTGCCCAGACAAGAGTCAA<br>TGTTCAGGTGGATGAGGTTCTCAGATCTAAGCACATGGGAGGGCGATCGCAATCTGGCTCCCAGTTTTGT<br>GAATGAAGATGGCGTCGAATGACGCCGCTCCATCTAATGATGGTGCTGCTGGTCTCGTACCAGAGGGCA<br>ACAACGAGACCCTTCCCCTAGAACCAGTTGCGGGCGCAGCTATAGCCGCACCCGTCACTGGCCAAAATA<br>ATGTAAT-3' | 629 out of 631 (99%) nucleotides match to KY905330.1 |
| GII-5 | 5'-<br>GACGGTGACTCGTGACCCAGCTGGCTGGTTTGGAAAACTGGACCAAAGTTCAATTTTGAGGCAGATGTA<br>CTGGACTAGAGGACCAAATCATGAAGACCCCAATGAGACAATGATACCCCATTCTCAAAGACCCATACA<br>GCTCATGGCACTGCTTGGTGAAGCCTCTCTTCACGGACCCTCTTTCTACAGTAGAATCAGTAAATTGGTC<br>ATAACTGAACTTAAAGAAGGTGGGATGGACTTTTACGTGCCAAGGCAGGAACCCATGTTCAGGTGGATG<br>AGGTTTTCTGACTTGAGCACGTGGGAGGGCGATCGCAATCTGGCTCCCAGCTTTGTGAATGAAGATGGCG<br>TCGAGTGACGCCAACCCATCTGATGGGTCCGCAGCCAACCTCGTACCAGAGGTCAACAATGAGGTTATG<br>GCTTTG-3' | 420 out of 422 (99%) nucleotides match to MK752943.1 |
| GII-6 | 5'-<br>CATACAGCTCATGGCACTGCTTGGTGAAGCCTCTCTTCACGGACCCTCTTTCTACAGTAGAATCAGCAAA<br>TTGGTCATAACTGGAACTTAAAGAAGGTGGTATGGATTTTTACGTGCCAAGACAGGAACCCATGTTCAG<br>GTGGATGAGGTTTTCTGACTTGAGCACGTGGGAGGGCGATCGCAATCTGGCTCCCAATTTTGTGAATGAA<br>GATGGCGTCGAGTGACGCCAACCCATCTGATGGGTCCGCAGCCAACCTCGTACCAGAGGTCAACAATGA<br>GGTTATGGCTTTGGAGCCCGTTGTTGGTGCCGCTATTGCGGCACCTGTAGCGGGCCAACAAAATGTAATT<br>GACCCCTGGATTAGAAATAATTTTGTACAAGCCCCTGGTGGGGAGTTTACAGTATCCCCTAGAAACGCTC<br>CAGGTGAAATACTATGGAGCGCGCCCCTAGGCCCCGACCTAAACCCCTATCTATCCCATTTGGCCAGAAT<br>GTACAATGGTTATGCAGGTGGTTTTGAAGTGCAGGTAATTCTCGCGGGGAACGCGTTCACCGCCGGGAA<br>GGTTATATTTGCAGCAGTCCCACCAAATTTTCCAACTGAAGGCTTAAGTCCTAGCCAGGTCACTATGTTC<br>CCCCATATAATAGTAGATGTTAGACAATTAGAACCTGTGCTAATTCCCTTACCCGATGTTAGGAATAATT<br>TTTATCATTACAATCAGTCAAATGACTCCACTATTAAGTTGATAGCAATGTTGTATACACCACTTAGGGC<br>TAATAATGCTGGGGATG-3' | 774 out of 784 (99%) nucleotides match to MW661260.1 |
| GII-7 | 5'-<br>CGGACCCTCTTTCTACAGTAGAATCAGCAAATTGGTCATAACTGAGCTCAAAGAAGGTGGGATGGACTTT<br>TACGTGCCAAGGCAGGAACCCATGTTCAGGTGGATGAGGTTTTCTGACTTGAGCACGTGGGAGGGCGAT<br>CGCAATCTGGCTCCCAATTTTGTGAATGAAGATGGCGTCGAATGACGCCAACCCATCTGATGGGTCCGCA<br>GCCAACCTCGTACCAGAGGTCAACAATGAGGTTATGGCTTTGGAGCCCGTTGTTGGTGCCGCTATTGCGG<br>CACCTGTAGCGGGCCAACAAAATGTAATTGACCCCTGGATTAGAAATAATTTTGTACAAGCCCCTGGCG<br>GGGAGTTCACAGTATCCCCTAGAAACGCTCCAGGTGAAATACTATGGAGCGCGCCCCTAGGCCCTGACC | 703 out of 712 (99%) nucleotides match to MW661278.1 |

| | | |
|---|---|---|
| | TAAATCCCTACCTGTCCCATTTGGCCAGAATGTACAATGGTTATGCAGGTGGTTTTGAAGTGCAGGTAAT<br>TCTCGCGGGGAACGCGTTCACCGCCGGGAAGATTATATTTGCAGCAGTCCCACCAAATTTTCCAACTGAA<br>GGCCTAAGTCCTAGCCAGGTCACTATGTTCCCCCACATAATAGTAGATGTTAGACAATTAGAACCTGTGC<br>TAATTCCCTTACCCGATGTTAGGAATAATTTCTATCATTACAATCAATCAAATGACTCCACTATTAAGTTG<br>ATAGCAATGTTGTA-3’ | |
| GII-8 | 5’-<br>ACAGTAGAATCAGCAAAATTGGTCATAACTGAGCTCAAAGAAGGTGGGATGGACTTTTACGTGCCAAGG<br>CAGGAACCCATGTTCAGGTGGATGAGGTTTTCTGACTTGAGCACGTGGGAGGGCGATCGCAATCTGGCT<br>CCCAATTTTGTGAATGAAGATGGCGTCGAATGACGCCAACCCATCTGATGGGTCCGCAGCCAACCTCGTA<br>CCAGAGGTCAACAATGAGGTTATGGCTTTGGAGCCCGTTGTTGGTGCCGCTATTGCGGCACCTGTAGCGG<br>GCCAACAAAATGTAATTGACCCCTGGATTAGAAATAATTTTGTACAAGCCCCTGGCGGGGAGTTCACAG<br>TATCCCCTAGAAACGCTCCAGGTGAAATACTATGGAGCGCGCCCCTAGGCCCTGACCTAAATCCCTACCT<br>GTCCCATTTGGCCAGAATGTACAATGGTTATGCAGGTGGTTTTGAAGTGCAGGTAATTCTCGCGGGGAAC<br>GCGTTCACCGCCGGGAAGATTATATTTGCAGCAGTCCCACCAAATTTTCCAACTGAAGGCCTAAGTCCTA<br>GCCAGGTCACTATGTTCCCCCACATAATAGTAGATGTTAGACAATTAGAACCTGTGCTAATTCCCTTACC<br>CGATGTTAGGAATAATTTCTATCATTACAATCAATCAAATGACTCCACTATTAAGTTGATAGCAATGTTG<br>T-3’ | 608 out of 615 (99%) nucleotides match to OM185499.1 |
| GII-9 | - | - |
| GII-10 | 5’-<br>CACCGACATAAAATTGGACCCTGAGCAGTTAACCGCCAAGTTGAGGGAGTACGGCCTGAAGCCAACCCG<br>CCCAGACAAGACCGAGGGACCCCTGATCATCAGTGAAGACTTGAACGGACTCACTTTCCTCCGAAGGAC<br>GGTGACTCGTGACCCAGCTGGCTGGTTTGGAAAACTGGATCAAAGTTCAATTCTGAGGCAGATGTACTG<br>GACTAGAGGACCAAATCATGAAGACCCCAATGAGACAATGATACCCCACTCTCAAAGACCCATACAGCT<br>CATGGCACTGCTTGGTGAAGCCTCTCTTCACGGACCCTCTTTCTACAGTAGAATCAGCAAATTGGTCATA<br>ACTGAGCTCAAAGAAGGTGGGATGGACTTTTACGTGCCAAGGCAGGAACCCATGTTCAGGTGGATGAGG<br>TTTTCTGACTTGAGCACGTGGGAGGGCGATCGCAATCTGGCTCCCAATTTTGTGAATGAAGATGGCGTCG<br>AATGACGCCAACCCATCTGATGGGTCCGCAGCCAACCTCGTACCAGAGGTCAACAATGAGGTTATGGCT<br>TTGGAGCCCGTTGTTGGTGCCGCTATTGCGGCACCTGTAGCGGGCCAACAAAATGTAATTG-3’ | 608 out of 615 (99%) nucleotides match to OM185499.1 |

1) Highlights indicate nucleotides that did not match to reference sequence. Underlines represent the annealing sites for three RT-qPCR assays (A1, A2, and A3 for GI samples and B1, B2, and B3 assay for GII samples).
2) The norovirus sequences were blasted to find the reference sequences (Genbank ID).

**Table S6.** The numbers in the right end column show the number of viral sequences with identical sequence to the PCR assay sequences / the total number of viral sequences from the database. The number in parenthesis shows the probability of detection in percentage.

| Target | Reference | type | 5’ to 3’ | In silico |
|---|---|---|---|---|
| RSV | (5) | Forward | CTTGATTCCTCGGTGTACCTCTGT | 28/1798 (2%) |
| | | Probe | TCCCATTAT/ZEN/GCCTAGGCCAGCAGCA | |
| | | Reverse | CTCAATTTCCTCACTTCTCCAGTGT | |
| | (6) | Forward | GCTCTTAGCAAAGTCAAGTTRAATGATACA | 422/1798 (23%) |
| | | Probe | CTGTCATCYAGCAAATACACTATCCAACGTAGCACAGG | |
| | | Reverse | GTTTTTGCACATCATAATTRGGAGT | |
| | (7) | Forward | CTCCAGAATAYAGGCATGAYTCTCC | 833/1798 (46%) |
| | | Probe | TAACCAAATTAGCAGCAGGAGATAGATCAG | |
| | | Reverse | GCYCTYCTAATYACWGCTGTAAGAC | |
| AdV | (1) | Forward | GCCTGGGGAACAAGTTCAGA | 126/126 (100%) |
| | | Probe | CAGTCGCTGYGACCTGTCTGTGGTT | |
| | | Reverse | GCGTAAAGCGCACTTTGTAAG | |
| | (8) | Forward | CCGACCCACGATGTAACCA | 54/126 (43%) |
| | | Probe | ACAGGTCACAGCGACTGACGCTGC | |
| | | Reverse | CGGTCGACTGGCACGAAT | |
| | (9) | Forward | TGGCCACCCCCTCGATGA | 97/126 (77%) |
| | | Probe | - | |
| | | Reverse | TTTAGGAGCCAGGGAGTTATA | |
| IAV | (10) | Forward | GACCRATCCTGTCACCTCTGAC | 2/11879 (0%) |
| | | Probe | TGCAGTCCTCGCTCACTGGGCACG | |
| | | Reverse | AGGGCATTYTGGACAAAKCGTCTA | |
| | (11) | Forward | CAAGACCAATCYTGTCACCTCTGAC | 101/1187<br>9<br>(1%) |
| | | Probe | TGCAGTCCTCGCTCACTGGGCACG | |
| | | Reverse | GCATTYTGGACAAAVCGTCTACG | |
| | (12) | Forward | CAAGACCAATCYTGTCACCTCTGAC<br>CAAGACCAATYCTGTCACCTYTGAC | 11094/11<br>879<br>(93%) |
| | | Probe | TGCAGTCCTCGCTCACTGGGCACG | |

| | | | | |
|---|---|---|---|---|
| | | Reverse | GCATTYTGGACAAAVCGTCTACG<br>GCATTTTGGATAAAGCGTCTACG | |
| RV | (13) | Forward | ATGTATGGTATTGAATATACC | 1216/1506<br>(81%)<br>vp7 |
| | | Probe | - | |
| | | Reverse | ACTTGCCACCATYTYTTCC | |
| | (14) | Forward | TCTGCAGACAGTTGAACCTATTAA,<br>CAGACACGGTTGAACCCATTAA,<br>TCGGCTGATACAGTAGAACCTATAAATG,<br>TGTCAGCTGATACAGTAGAACCTATAAATG,<br>TCAGCTGACACAGTAGAACCTATAAATG | 322/2012<br>(16%)<br>vp2 |
| | | Probe | ATGCGCATRTTRTCAAAHGCAA | |
| | | Reverse | GTTGGCGTTTACAGTTCGTTCAT,<br>GTTGGCGTCTACAATTCGTTCAT | |
| | (15) | Forward | GTNTTTCCACCAGGYATGA | 8/257<br>(3%)<br>vp6 |
| | | Probe | GGTCACATCCTCTCACTA | |
| | | Reverse | CACCATCHAGRGARGATAA | |

### Table S7. Interaction between A1 assay and GI clinical samples

| Sample | Forward primer | Probe | Reverse primer |
|---|---|---|---|
| GI-1 | - | - | - |
| GI-2 | 5’-GCHATRTTYCGYTGGATG-3’<br>5’-GCCATGTTCCGCTGGATG-3’ | 5’-TGGACAGGRGAYCGCRATCT-3’<br>5’-TGGACAGGGGACCGCGATCT-3’ | 5’-CTTAGACGCCATCATCATT-3’<br>5’-CTTAGACGCCATCATCATT-3’(R) |
| GI-3 | 5’-GCHATRTTYCGYTGGATG-3’<br>5’-GCCATGTTCCGCTGGATG-3’ | 5’-TGGACAGGRGAYCGCRATCT-3’<br>5’-TGGACAGGAGACCGCGATCT-3’ | 5’-CTTAGACGCCATCATCATT-3’<br>5’-CTTAGACGCCATCATCATT-3’(R) |
| GI-4 | 5’-GCHATRTTYCGYTGGATG-3’<br>5’-GCCATATTCCGCTGGATG-3’ | 5’-TGGACAGGRGAYCGCRATCT-3’<br>5’-TGGACAGGAGATCGCAATCT-3’ | 5’-CTTAGACGCCATCATCATT-3’<br>5’-CTTAGACGCCATCATCATT-3’(R) |
| GI-5 | 5’-GCHATRTTYCGYTGGATG-3’<br>5’-GCCATATTCCGCTGGATG-3’ | 5’-TGGACAGGRGAYCGCRATCT-3’<br>5’-TGGACAGGAGATCGCAATCT-3’ | 5’-CTTAGACGCCATCATCATT-3’<br>5’-CTTAGACGCCATCATCATT-3’(R) |
| GI-6 | 5’-GCHATRTTYCGYTGGATG-3’<br>5’-GCCATATTCCGCTGGATG-3’ | 5’-TGGACAGGRGAYCGCRATCT-3’<br>5’-TGGACAGGAGATCGCAATCT-3’ | 5’-CTTAGACGCCATCATCATT-3’<br>5’-CTTAGACGCCATCATCATT-3’(R) |
| GI-7 | - | - | - |
| GI-8 | 5’-GCHATRTTYCGYTGGATG-3’<br>5’-GCCATATTCCGCTGGATG-3’ | 5’-TGGACAGGRGAYCGCRATCT-3’<br>5’-TGGACAGGAGATCGCAATCT-3’ | 5’-CTTAGACGCCATCATCATT-3’<br>5’-CTTAGACGCCATCATCATT-3’(R) |
| GI-9 | - | - | - |
| GI-10 | 5’-GCHATRTTYCGYTGGATG-3’<br>5’-GCCATGTTCCGCTGGATG-3’ | 5’-TGGACAGGRGAYCGCRATCT-3’<br>5’-TGGACAGGAGACCGCGATCT-3’ | 5’-CTTAGACGCCATCATCATT-3’<br>5’-CTTAGACGCCATCATCATT-3’(R) |

### Table S8. Interaction between A2 assay and GI clinical samples

| Sample | Forward primer | Probe | Reverse primer |
|---|---|---|---|
| GI-1 | - | - | - |
| GI-2 | 5’-GCYATGTTCCGYTGGATGC-3’<br>5’-GCCATGTTCCGCTGGATGC-3’ | 5’-TCGGGCAGGAGATYGCGRTCYC-3’<br>5’-TCGGGCAGGAGATCGCGGTCCC-3’(R) | 5’-GTCCTTAGACGCCATCATCATT-3’<br>5’-GTCCTTAGACGCCATCATCATT-3’(R) |
| GI-3 | 5’-GCYATGTTCCGYTGGATGC-3’<br>5’-GCCATGTTCCGCTGGATGC-3’ | 5’-TCGGGCAGGAGATYGCGRTCYC-3’<br>5’-TCGGGCAAGAGATCGCGGTCTC-3’(R) | 5’-GTCCTTAGACGCCATCATCATT-3’<br>5’-GTCCTTAGACGCCATCATCATT-3’(R) |
| GI-4 | 5’-GCYATGTTCCGYTGGATGC-3’<br>5’-GCCATATTCCGCTGGATGC-3’ | 5’-TCGGGCAGGAGATYGCGRTCYC-3’<br>5’-TCGGGCAGGAGATTGCGATCTC-3’(R) | 5’-GTCCTTAGACGCCATCATCATT-3’<br>5’-GTCCTTAGACGCCATCATCATT-3’(R) |
| GI-5 | 5’-GCYATGTTCCGYTGGATGC-3’<br>5’-GCCATATTCCGCTGGATGC-3’ | 5’-TCGGGCAGGAGATYGCGRTCYC-3’<br>5’-TCGGGCAGGAGATTGCGATCTC-3’(R) | 5’-GTCCTTAGACGCCATCATCATT-3’<br>5’-GTCCTTAGACGCCATCATCATT-3’(R) |
| GI-6 | 5’-GCYATGTTCCGYTGGATGC-3’<br>5’-GCCATATTCCGCTGGATGC-3’ | 5’-TCGGGCAGGAGATYGCGRTCYC-3’<br>5’-TCGGGCAGGAGATTGCGATCTC-3’(R) | 5’-GTCCTTAGACGCCATCATCATT-3’<br>5’-GTCCTTAGACGCCATCATCATT-3’(R) |
| GI-7 | - | - | - |
| GI-8 | 5’-GCYATGTTCCGYTGGATGC-3’<br>5’-GCCATATTCCGCTGGATGC-3’ | 5’-TCGGGCAGGAGATYGCGRTCYC-3’<br>5’-TCGGGCAGGAGATTGCGATCTC-3’(R) | 5’-GTCCTTAGACGCCATCATCATT-3’<br>5’-GTCCTTAGACGCCATCATCATT-3’(R) |
| GI-9 | - | - | - |
| GI-10 | 5’-GCYATGTTCCGYTGGATGC-3’<br>5’-GCCATGTTCCGCTGGATGC-3’ | 5’-TCGGGCAGGAGATYGCGRTCYC-3’<br>5’-TCGGGCAAGAGATCGCGGTCTC-3’(R) | 5’-GTCCTTAGACGCCATCATCATT-3’<br>5’-GTCCTTAGACGCCATCATCATT-3’(R) |

### Table S9. Interaction between A3 assay and GI clinical samples

| Sample | Forward primer | Probe | Reverse primer |
|---|---|---|---|
| GI-1 | - | - | - |
| GI-2 | 5’-GGAGATCGCRATCTCCTGCC-3’<br>5’-GGGGACCGCGATCTCCTGCC-3’ | 5’-TAAATGATGATGGCGTCTAAGGACGCCCC-3’<br>5’-TAAATGATGATGGCGTCTAAGGACGCCCC-3’ | 5’-CTCYGGTACCAGCTGGCC-3’<br>5’-CTCTGGTACCAGCTGGCC-3’(R) |
| GI-3 | 5’-GGAGATCGCRATCTCCTGCC-3’<br>5’-GGAGACCGCGATCTCTTGCC-3’ | 5’-TAAATGATGATGGCGTCTAAGGACGCCCC-3’<br>5’-TAAATGATGATGGCGTCTAAGGACGCCCC-3’ | 5’-CTCYGGTACCAGCTGGCC-3’<br>5’-CTCTGGTACCAGCTGACC-3’(R) |
| GI-4 | 5’-GGAGATCGCRATCTCCTGCC-3’<br>5’-GGAGATCGCAATCTCCTGCC-3’ | 5’-TAAATGATGATGGCGTCTAAGGACGCCCC-3’<br>5’-TAAATGATGATGGCGTCTAAGGACGCCCC-3’ | 5’-CTCYGGTACCAGCTGGCC-3’<br>5’-CTCTGGAACCAGCTGACC-3’(R) |
| GI-5 | 5’-GGAGATCGCRATCTCCTGCC-3’<br>5’-GGAGATCGCAATCTCCTGCC-3’ | 5’-TAAATGATGATGGCGTCTAAGGACGCCCC-3’<br>5’-TAAATGATGATGGCGTCTAAGGACGCCCC-3’ | 5’-CTCYGGTACCAGCTGGCC-3’<br>5’-CTCTGGAACCAGCTGACC-3’(R) |
| GI-6 | 5’-GGAGATCGCRATCTCCTGCC-3’<br>5’-GGAGATCGCAATCTCCTGCC-3’ | 5’-TAAATGATGATGGCGTCTAAGGACGCCCC-3’<br>5’-TAAATGATGATGGCGTCTAAGGACGCCCC-3’ | 5’-CTCYGGTACCAGCTGGCC-3’<br>5’-CTCTGGAACCAGCTGACC-3’(R) |
| GI-7 | - | - | - |
| GI-8 | 5’-GGAGATCGCRATCTCCTGCC-3’<br>5’-GGAGATCGCAATCTCCTGCC-3’ | 5’-TAAATGATGATGGCGTCTAAGGACGCCCC-3’<br>5’-TAAATGATGATGGCGTCTAAGGACGCCCC-3’ | 5’-CTCYGGTACCAGCTGGCC-3’<br>5’-CTCTGGAACCAGCTGACC-3’(R) |
| GI-9 | - | - | - |
| GI-10 | 5’-GGAGATCGCRATCTCCTGCC-3’<br>5’-GGAGACCGCGATCTCTTGCC-3’ | 5’-TAAATGATGATGGCGTCTAAGGACGCCCC-3’<br>5’-TAAATGATGATGGCGTCTAAGGACGCCCC-3’ | 5’-CTCYGGTACCAGCTGGCC-3’<br>5’-CTCTGGTACCAGCTGACC-3’(R) |

### Table S10. Interaction between B1 assay and GII clinical samples

| Sample | Forward primer | Probe | Reverse primer |
|---|---|---|---|
| GII-1 | 5’-TTYAGRTGGATGAGRTTYTC-3’<br>5’-TTTAGATGGATGAGATTTTC-3’ | 5’-ACDTGGGAGGGYGATCRCAAT-3’<br>5’-ACGTGGGAGGGCGATCGCAAT-3’ | 5’-YMGAYGCCATCHTCATTC-3’<br>5’-GAATGAAGATGGCGTCGA-3’(R) |
| GII-2 | - | - | - |
| GII-3 | 5’-TTYAGRTGGATGAGRTTYTC-3’<br>5’-TTCAGGTGGATGAGGTTCTC-3’ | 5’-ACDTGGGAGGGYGATCRCAAT-3’<br>5’-ACATGGGAGGGCGATCGCAAT-3’ | 5’-YMGAYGCCATCHTCATTC-3’<br>5’-TCGACGCCATCTTCATTC-3’(R) |
| GII-4 | 5’-TTYAGRTGGATGAGRTTYTC-3’<br>5’-TTCAGGTGGATGAGGTTCTC-3’ | 5’-ACDTGGGAGGGYGATCRCAAT-3’<br>5’-ACATGGGAGGGCGATCGCAAT-3’ | 5’-YMGAYGCCATCHTCATTC-3’<br>5’-TCGACGCCATCTTCATTC-3’(R) |
| GII-5 | 5’-TTYAGRTGGATGAGRTTYTC-3’<br>5’-TTCAGGTGGATGAGGTTTTC-3’ | 5’-ACDTGGGAGGGYGATCRCAAT-3’<br>5’-ACGTGGGAGGGCGATCGCAAT-3’ | 5’-YMGAYGCCATCHTCATTC-3’<br>5’-TCGACGCCATCTTCATTC-3’(R) |
| GII-6 | 5’-TTYAGRTGGATGAGRTTYTC-3’<br>5’-TTCAGGTGGATGAGGTTTTC-3’ | 5’-ACDTGGGAGGGYGATCRCAAT-3’<br>5’-ACGTGGGAGGGCGATCGCAAT-3’ | 5’-YMGAYGCCATCHTCATTC-3’<br>5’-TCGACGCCATCTTCATTC-3’(R) |
| GII-7 | 5’-TTYAGRTGGATGAGRTTYTC-3’<br>5’-TTCAGGTGGATGAGGTTTTC-3’ | 5’-ACDTGGGAGGGYGATCRCAAT-3’<br>5’-ACGTGGGAGGGCGATCGCAAT-3’ | 5’-YMGAYGCCATCHTCATTC-3’<br>5’-TCGACGCCATCTTCATTC-3’(R) |
| GII-8 | 5’-TTYAGRTGGATGAGRTTYTC-3’<br>5’-TTCAGGTGGATGAGGTTTTC-3’ | 5’-ACDTGGGAGGGYGATCRCAAT-3’<br>5’-ACGTGGGAGGGCGATCGCAAT-3’ | 5’-YMGAYGCCATCHTCATTC-3’<br>5’-TCGACGCCATCTTCATTC-3’(R) |
| GII-9 | - | - | - |
| GII-10 | 5’-TTYAGRTGGATGAGRTTYTC-3’<br>5’-TTCAGGTGGATGAGGTTTTC-3’ | 5’-ACDTGGGAGGGYGATCRCAAT-3’<br>5’-ACGTGGGAGGGCGATCGCAAT-3’ | 5’-YMGAYGCCATCHTCATTC-3’<br>5’-TCGACGCCATCTTCATTC-3’(R) |

### Table S11. Interaction between B2 assay and GII clinical samples

| Sample | Forward primer | Probe | Reverse primer |
|---|---|---|---|
| GII-1 | 5’-ATGTTCAGRTGGATGAGRTTCTCWGA-3’<br>5’-ATGTTTAGATGGATGAGATTTTCAGA-3’ | 5’-AGCACGTGGGAGGGCGATCG-3’<br>5’-AGCACGTGGGAGGGCGATCG-3’ | 5’-TCGACGCCATCTTCATTCACA-3’<br>5’-TGTGAATGAAGATGGCGTCGA-3’(R) |
| GII-2 | - | - | - |
| GII-3 | 5’-ATGTTCAGRTGGATGAGRTTCTCWGA-3’<br>5’-ATGTTCAGGTGGATGAGGTTCTCAGA-3’ | 5’-AGCACGTGGGAGGGCGATCG-3’<br>5’-AGCACATGGGAGGGCGATCG-3’ | 5’-TCGACGCCATCTTCATTCACA-3’<br>5’-TCGACGCCATCTTCATTCACA-3’(R) |
| GII-4 | 5’-ATGTTCAGRTGGATGAGRTTCTCWGA-3’<br>5’-ATGTTCAGGTGGATGAGGTTCTCAGA-3’ | 5’-AGCACGTGGGAGGGCGATCG-3’<br>5’-AGCACATGGGAGGGCGATCG-3’ | 5’-TCGACGCCATCTTCATTCACA-3’<br>5’-TCGACGCCATCTTCATTCACA-3’(R) |
| GII-5 | 5’-ATGTTCAGRTGGATGAGRTTCTCWGA-3’<br>5’-ATGTTCAGGTGGATGAGGTTTTCTGA-3’ | 5’-AGCACGTGGGAGGGCGATCG-3’<br>5’-AGCACGTGGGAGGGCGATCG-3’ | 5’-TCGACGCCATCTTCATTCACA-3’<br>5’-TCGACGCCATCTTCATTCACA-3’(R) |
| GII-6 | 5’-ATGTTCAGRTGGATGAGRTTCTCWGA-3’<br>5’-ATGTTCAGGTGGATGAGGTTTTCTGA-3’ | 5’-AGCACGTGGGAGGGCGATCG-3’<br>5’-AGCACGTGGGAGGGCGATCG-3’ | 5’-TCGACGCCATCTTCATTCACA-3’<br>5’-TCGACGCCATCTTCATTCACA-3’(R) |
| GII-7 | 5’-ATGTTCAGRTGGATGAGRTTCTCWGA-3’<br>5’-ATGTTCAGGTGGATGAGGTTTTCTGA-3’ | 5’-AGCACGTGGGAGGGCGATCG-3’<br>5’-AGCACGTGGGAGGGCGATCG-3’ | 5’-TCGACGCCATCTTCATTCACA-3’<br>5’-TCGACGCCATCTTCATTCACA-3’(R) |
| GII-8 | 5’-ATGTTCAGRTGGATGAGRTTCTCWGA-3’<br>5’-ATGTTCAGGTGGATGAGGTTTTCTGA-3’ | 5’-AGCACGTGGGAGGGCGATCG-3’<br>5’-AGCACGTGGGAGGGCGATCG-3’ | 5’-TCGACGCCATCTTCATTCACA-3’<br>5’-TCGACGCCATCTTCATTCACA-3’(R) |
| GII-9 | - | - | - |
| GII-10 | 5’-ATGTTCAGRTGGATGAGRTTCTCWGA-3’<br>5’-ATGTTCAGGTGGATGAGGTTTTCTGA-3’ | 5’-AGCACGTGGGAGGGCGATCG-3’<br>5’-AGCACGTGGGAGGGCGATCG-3’ | 5’-TCGACGCCATCTTCATTCACA-3’<br>5’-TCGACGCCATCTTCATTCACA-3’(R) |

### Table S12. Interaction between B3 assay and GII clinical samples

| Sample | Forward primer | Probe | Reverse primer |
|---|---|---|---|
| GII-1 | 5’-GTGGGATGGACTTTTACGTGCCAA-3’<br>5’-GTGGTCTGGAGTTTTACGTGCCCA-3’ | 5’-AGCCAGATTGCGATCGCCCTCC-3’<br>5’-AGCCAGATTGCGATCGCCCTCC-3’(R) | 5’-CGTCAYTCGACGCCATCTTCATTCAC-3’<br>5’-CGTCATTCGACGCCATCTTCATTCAC-3’(R) |
| GII-2 | - | - | - |
| GII-3 | 5’-GTG GGATGGACTTTTACGTGCCAA-3’<br>5’-CTGAGGATGGAGTTTT GGTCCAA-3’ | 5’-AGCCAGATTGCGATCGCCCTCC-3’<br>5’-AGCCAGATTGCGATCGCCCTCC-3’(R) | 5’-CGTCAYTCGACGCCATCTTCATTCAC-3’<br>5’-CGTCATTCGACGCCATCTTCATTCAC-3’(R) |
| GII-4 | 5’-GTG GGATGGACTTTTACGTGCCAA-3’<br>5’-CTGAGGATGGAGTTTT GGTCCAA-3’ | 5’-AGCCAGATTGCGATCGCCCTCC-3’<br>5’-AGCCAGATTGCGATCGCCCTCC-3’(R) | 5’-CGTCAYTCGACGCCATCTTCATTCAC-3’<br>5’-CGTCATTCGACGCCATCTTCATTCAC-3’(R) |
| GII-5 | 5’-GTGGGATGGACTTTTACGTGCCAA-3’<br>5’-GTGGGATGGACTTTTACGTGCCAA-3’ | 5’-AGCCAGATTGCGATCGCCCTCC-3’<br>5’-AGCCAGATTGCGATCGCCCTCC-3’(R) | 5’-CGTCAYTCGACGCCATCTTCATTCAC-3’<br>5’-CGTCACTCGACGCCATCTTCATTCAC-3’(R) |
| GII-6 | 5’-GTGGGATGGACTTTTACGTGCCAA-3’<br>5’-GTGGTATGGATTTTTACGTGCCAA-3’ | 5’-AGCCAGATTGCGATCGCCCTCC-3’<br>5’-AGCCAGATTGCGATCGCCCTCC-3’(R) | 5’-CGTCAYTCGACGCCATCTTCATTCAC-3’<br>5’-CGTCACTCGACGCCATCTTCATTCAC-3’(R) |
| GII-7 | 5’-GTGGGATGGACTTTTACGTGCCAA-3’<br>5’-GTGGGATGGACTTTTACGTGCCAA-3’ | 5’-AGCCAGATTGCGATCGCCCTCC-3’<br>5’-AGCCAGATTGCGATCGCCCTCC-3’(R) | 5’-CGTCAYTCGACGCCATCTTCATTCAC-3’<br>5’-CGTCATTCGACGCCATCTTCATTCAC-3’(R) |
| GII-8 | 5’-GTGGGATGGACTTTTACGTGCCAA-3’<br>5’-GTGGGATGGACTTTTACGTGCCAA-3’ | 5’-AGCCAGATTGCGATCGCCCTCC-3’<br>5’-AGCCAGATTGCGATCGCCCTCC-3’(R) | 5’-CGTCAYTCGACGCCATCTTCATTCAC-3’<br>5’-CGTCATTCGACGCCATCTTCATTCAC-3’(R) |
| GII-9 | - | - | - |
| GII-10 | 5’-GTGGGATGGACTTTTACGTGCCAA-3’<br>5’-GTGGGATGGACTTTTACGTGCCAA-3’ | 5’-AGCCAGATTGCGATCGCCCTCC-3’<br>5’-AGCCAGATTGCGATCGCCCTCC-3’(R) | 5’-CGTCAYTCGACGCCATCTTCATTCAC-3’<br>5’-CGTCATTCGACGCCATCTTCATTCAC-3’(R) |

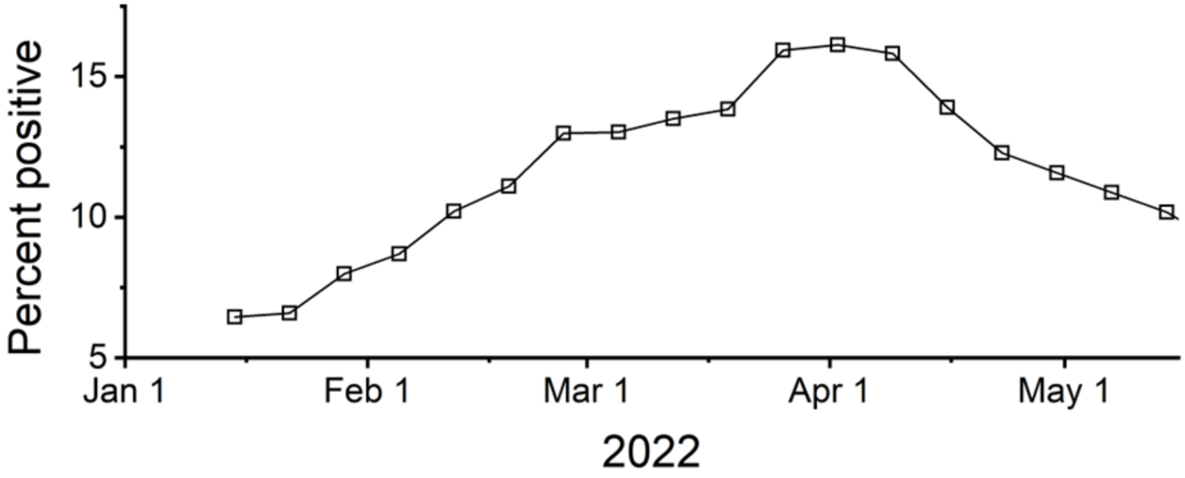


**Fig. S5**. Percent positive of residents in Midwestern United States for norovirus determined by PCR test. This norovirus surveillance data was published by the National Respiratory and Enteric Virus Surveillance System (NREVSS) and the raw data for this graph was downloaded from https://www.cdc.gov/surveillance/nrevss/norovirus/region.html.

**Table S13.** Summary of previous reports on evolutionary rate and time to the most recent common ancestor (TMRCA) of ten viral species

| Genome type | Viral species | Evolutionary rate | TMRCA | Note | Reference |
|---|---|---|---|---|---|
| RNA | NV | $1.2\times10^{-2}$ | 1999 | VP1 gene of GII.4 genotype | (16) |
| | RSV | $1.5\times10^{-3}$ s/s/y | 1943 | RSV-A | (17) |
| | Rotavirus | $1.9\times10^{-3}$ | 1989 | G9 type | (18) |
| | Influenza A virus | $2.9\times10^{-3}$ | 1898 | H1N1 strain | (19) |
| | SARS-CoV-2 | $6.7\times10^{-4}$ to $3.3\times10^{-3}$ | 2019 | - | (20–22) |
| | SARS-CoV-1 | $4\times10^{-4}$ | 2002 | - | (23) |
| | MERS | $1.1\times10^{-3}$ | 2012 | - | (24) |
| | Ebola | $1.2\times10^{-3}$ | 2013 | | (25) |
| DNA | AdV | $4.1\times10^{-5}$ | 1720 | Type 41 | (26) |
| | Mpox | $5\times10^{-6}$ | 2022 | orthopoxviruses | (27) |
